# The impact of opening dedicated clinics on disease transmission during an influenza pandemic

**DOI:** 10.1101/2020.05.13.20099622

**Authors:** Pengyi Shi, Jia Yan, Pinar Keskinocak, Andi L. Shane, Julie L. Swann

**Affiliations:** Krannert School of Management, Purdue University, West Lafayette, Indiana, United States of America; School of Industrial and Systems Engineering, Georgia Institute of Technology, Atlanta, Georgia, United States of America; Division of Infectious Diseases, Department of Pediatrics, Emory University and Children’s Healthcare of Atlanta, Atlanta, Georgia, United States of America; Department of Industrial and Systems Engineering, North Carolina State University, Raleigh, North Carolina, United States of America

**Keywords:** Pandemics, Human Influenza, Theoretical Models, Prevalence, Masks, Health Facilities

## Abstract

Dedicated clinics can be established in an influenza pandemic to isolate people and potentially reduce opportunities for influenza transmission. However, their operation requires resources and their existence may attract the worried-well. In this study, we quantify the impact of opening dedicated influenza clinics during a pandemic based on an agent-based simulation model across a time-varying social network of households, workplaces, schools, community locations, and health facilities in the state of Georgia. We calculate performance measures, including peak prevalence and total attack rate, while accounting for clinic operations, including timing and location. We find that opening clinics can reduce disease spread and hospitalizations even when visited by the worried-well, open for limited weeks, or open in limited locations, and especially when the clinics are in operation during times of highest prevalence. Specifically, peak prevalence, total attack rate, and hospitalization reduced 0.07-0.32%, 0.40-1.51%, 0.02-0.09%, respectively, by operating clinics for the pandemic duration.

## Introduction

During the H1N1 influenza pandemic in 2009-2020, many people visited health facilities to seek diagnoses and treatment [1]. Visits to emergency departments (EDs) surge, which might result in opportunities for transmission to others. As a result, some facilities chose to dedicate space and resources to the establishment of clinics, which could diagnose and manage people with known or suspected influenza infections to divert them from EDs [2–4]. Dedicated influenza clinics could help to separate people with influenza-like illness (“ILI patients”) from other people seeking care for a non-ILI diagnosis (“non-ILI patients”), and thus reduce transmission to uninfected people who had the potential for a severe ILI manifestation if exposed. However, dedicated influenza clinics required human and material resources at a time when a system would be operating at full capacity. Additionally, dedicated influenza clinics could attract the worried-well, that is people who do not have the flu but are worried enough that they visit the flu clinic to be sure, seeking reassurance, utilizing resources, and potentially exposing themselves to others [5].

Two recent observational studies emphasized the importance of dedicated clinics during an influenza pandemic. FitzGerald et al. [6] reviewed the impact of the 2009 H1N1 influenza on ED operation in Australia. They concluded that dedicated influenza clinics could help manage people in an influenza pandemic and noted the importance of personal protective equipment and antivirals therapy in disease management. An observational retrospective study [7] in Taiwan found that a dedicated influenza clinic external to an ED could reduce the length of stay compared to regular ED services. However, both studies were observational, so it is difficult to quantify the impact of dedicated clinics during an influenza pandemic under different scenarios, project the resources needed, or compare dedicated clinics to other interventions.

In this study, we utilized an agent-based simulation to evaluate the impact of dedicated influenza clinics functioning for the duration of the pandemic versus for a limited time. We evaluated the changes in the prevalence of infection, the total attack rate in population at risk, hospitalizations, and transmission of infections in hospitals along with the resources needed to operate the clinics for different periods of time. Agent-based simulations have been widely used to model the spread of influenza in prior studies [8–13]; however, these models have not captured disease transmission occurring specifically in health facilities. A key feature of our study is that dedicated clinics may take time before they can be open, and they may not be open throughout the disease spread or across all locations. We accounted for people at higher risk of developing flu-related complications, e.g., young children, the elderly, pregnant women, and people with existing medical conditions [14], who seek healthcare at greater rates than lower-risk people. We also allowed for influenza clinics to bring together people who have influenza-like illnesses but may or may not have the flu, denoted at the “worried-well”. We compared the impact of dedicated influenza clinics with and without extensive use of masks in health facilities. We modeled disease spread in households, workplaces and schools, and the community among census tracts and counties in the state of Georgia [15] and quantified the impact of dedicated influenza clinics on transmission.

## Methods

Our agent-based simulation model included two critical components: (i) the disease progression within each agent (individual) and (ii) the contact network. Each agent in our model corresponded to an individual with certain social and geographical characteristics. The full details of the model (e.g., specifics on mixing, transmission, contact networks, etc.) are available in the Appendix.

### (i) Disease progression

The progression of flu within an individual is described using a Susceptible-Exposed-Infectious-Recovered (SEIR) model [16–18]. We described the progression of influenza with a refined SEIR model [19–22], which divided the infectious stage into more detailed sub-stages. Each agent was assumed to be in one of the following states: susceptible (*S*), exposed (*E*), infectious and presymptomatic (*I_P_*), infectious and symptomatic (*I_S_*), infectious and asymptomatic (*I_A_*), infectious and hospitalized (*I_H_*), recovered (*R*) or dead (*D*). All agents started in the susceptible state. The transition diagram is found in [22]. Agents are classified according to five age groups: 0-5, 6-11, 12-18, 19-64, and 65+ years. We assumed high-risk agents, e.g., people with co-morbidities that made them more vulnerable to severe outcomes from the flu, had a higher frequency of healthcare visits, and the probability of being hospitalized if influenza was contracted than low-risk ones. Hospitalizations are considered a severe outcome of influenza, which is typically associated with high mortality for the patients and a longer duration of being infectious to their contacts. The age- and risk-level specific transition probabilities and duration in each (sub-)stage are in Table 1. As the number of high-risk individuals may not be known, we established lower and upper bounds ([LB, UB]) for children [12%, 24%] and adults [8%, 24%] who were likely in this category. The details of the estimation are presented in the Appendix.

**Table 1.** Notations of constants used in our agent-based simulation model.

| Parameter | Description | Values | References |
| --- | --- | --- | --- |
| $p_A$ | Probability of a presymptomatic person becoming asymptomatic | 0.4 for adults aged 19-64, 0.25 for others | [22-25] |
| $p_H$ | Probability of a symptomatic person being hospitalized | 0.18 for low-risk young children aged 0-5, 0.06 for low-risk people aged 6-64, 0.12 for adults aged above 65<br>0.36 for high-risk children aged 0-5, 0.12 for high-risk children aged 6-18, other high-risk age groups are the same as low-risk people | [22, 25] |
| $p_D$ | Probability of death for a hospitalized patient | 0.344 for young children aged 0-5, and adults aged above 65, 0.172 for others | [22, 26] |
| Duration of $E+I_P$ | Duration of exposed and presymptomatic stages | Weibull distribution with mean 1.48 days (including an offset of 0.5 days) and standard deviation 0.47 days | [22, 27] |
| Duration of $I_P$ | Duration of presymptomatic stage | 0.5 days | [22, 27] |
| Duration of $I_S$ | Duration of symptomatic stage | Exponential distribution with mean 2.73 days | [22] |
| Duration of $I_A$ | Duration of asymptomatic stage | Exponential distribution with mean 1.64 days | [22] |
| Duration of $I_H$ | Duration of hospitalization stage | Exponential distribution with mean 14 days | [22, 27] |
| Household size | Distribution of number of agents in one household | 1-person household: 10.33%;<br>2-person household: 23.55%;<br>3-person household: 20.45%;<br>4-person household: 23%;<br>5-person household: 12.79%;<br>6-person household: 5.91%;<br>7-person household: 3.97%. | [15] |
| Classroom size | Distribution of number of students in one classroom | Age 0-5: uniform distribution (9,19); 6-11: uniform distribution (15,25); 12-18: uniform (25,35). | [22, 28] |
| Workplace size | Distribution of number of staff in one workplace | Poisson distribution with mean 20 (upper bound 1000) | [22, 25] |
| Community size | Number of agents in each census tract | Values from data source; maximum: 29341, minimum: 218 | [15] |
| $q_n$ | Frequency of healthcare encounters for non-ILI<sup>a</sup> patients | 1 per year for low-risk non-ILI<sup>a</sup> patients, 4 per year for high-risk non-ILI<sup>a</sup> patients | [29] |

<sup>a</sup> ILI: influenza-like illness

We also assumed that anyone who recovers from influenza during the time horizon of the model is recovered with immunity and cannot infect others.

This model has been validated against previous pandemics, and versions of it have been published in several other papers (See Appendix).

### (ii) Contact network

Agents could contact each other within their social groups, including household (*H*), community (*C*), peer groups (*G*), hospital (*D*), and dedicated influenza clinic (*F*) if open. The hospital consists of an emergency department (ED) for short-term acute care and impatients who are admitted (or hospitalized) for care for at least one night. The peer group refers to schools or workplaces (based on the age group of each agent), and the community group is used to capture random contacts such as in churches or stores [19, 21]. The size distribution of social groups is in Table 1.

The model represented agents at the level of census tracts, with each assigned to a household size based on census data for the tract. During the daytime, agents are assigned to schools (including daycare, preschools), workplaces, or by themselves based on their age groups (≤18, 19-64, 65+ years old, respectively). All agents interact within their household at night and in communities (e.g., grocery stores) during both day and night. If a young patient (≤18 years old) is symptomatic, the person will withdraw from school; if an adult patient is symptomatic, the person will withdraw from work with a probability of 0.5.

We assume each agent is associated with the closest of 152 short-term acute care, critical access (e.g., providing healthcare for common conditions in rural areas), or children’s hospitals in Georgia, and each hospital could establish up to one dedicated influenza clinic to serve individuals associated with the hospital. We acknowledge that some patients may present to general practitioners. We are focused here on cases that need a great level of care, or on patients sent by general practitioners. Health facilities might are by agents in two categories: *ILI patients* who visited health facilities because they showed ILI symptoms, and *non-ILI patients* who sought care for diagnoses other than ILI. ILI-patients included those who were infected (flu patients) plus some worried-well individuals who thought they might be infected. We assumed worried-wells were present only when dedicated influenza clinics were open and that the number of the worried-well was proportional to the number of flu patients in the same clinic on that day. *P_ww_* denotes the ratio of worried-well to other ILI patients; values are shown in Table 2. The timing of care seeking for patients who have influenza is random within the period where they are infectious and showing symptoms. Patients who have influenza are hospitalized according to the disease progression and can be admitted for overnight stays from clinics, EDs, or from the community.

**Table 2.** Notation and levels of parameters for baseline and sensitivity analysis.

| Notation | Description | Levels of parameters |
| --- | --- | --- |
| Clinic switch | Switch to settings with/out clinics | • NC (baseline): no clinic<br>• CL: opening clinics based on $T_c$, $X_c$, and $L_c$ |
| $MX$ | Mixing mode at night | • FM (baseline): mixing with family at night<br>• PT: mixing with other patients at night |
| $P_{hr}$ | Proportion of high-risk people | • UB (baseline): 22% for the children and 24% for adults<br>• LB: 12% for the children and 8% for adults<br>Estimated based on (22-23), details in the Appendix |
| $v_f$ | Probability to visit hospitals and clinics for flu patients | • Medium (baseline): 25% for low-risk flu patients and 50% for high-risk flu patients if clinic is not open, and 50% for low-risk flu patients and 100% for high-risk flu patients if clinic is open<br>• Low: 10% for low-risk flu patients and 20% for high-risk flu patients if clinic is not open, and 20% for low-risk flu patients and 40% for high-risk flu patients if clinic is open<br>• High: 37.5% for low-risk flu patients and 75% for high-risk flu patients if clinic is not open, and 75% for low-risk flu patients and 100% for high-risk flu patients if clinic is open<br>Estimated from [30] |
| $q_f$ | Frequency to visit hospitals and clinics for flu patients | $v_f$ /duration of $I_s$<br>• Medium (baseline), low and high values based on $v_f$ |
| $T_c$ | Initiation of operation date of clinic C | • Week 1 (baseline)<br>• Week 4<br>• Week 5<br>• Week 6<br>• Week 7<br>• Week 8<br>• Week 9<br>• Week 10 |
| $X_c$ | Durations of clinic C | • 1 year (baseline)<br>• 4 weeks<br>• 8 weeks |
| $L_c$ | Location of clinic C | • Location Group 1 - metropolitan Atlanta (Cherokee, Clayton, Cobb, DeKalb, Douglas, Fayette, Fulton, Gwinnett, Henry, Rockdale counties)<br>• Location Group 2 – other locations |
| Mask types | Different masks have different effects on susceptibility and infectivity | • N95 (baseline): decreasing susceptibility by 20% and infectivity by 50%[31]<br>• Surgical mask: decreasing susceptibility by 2% and infectivity by 5% [31] |
| $P_{mk}$ | Percentage of people wearing masks in hospitals and clinics | • No mask (baseline): 0%<br>• 25%<br>• 50%<br>• 100% |
| Initial $R_0$ | Reproductive rate defined as average number of secondary cases generated by each infected patient before interventions are introduced | • 1.5 (baseline)<br>• 1.8<br>[22, 25, 27, 32, 33] |
| $P_{ww}$ | Number of worried-well over number of flu patients in the same clinic | • 0.5 (baseline)<br>• 0.2 |

The interactions (or lack thereof) between ILI and non-ILI patients is partly determined by the time of day and whether a dedicated influenza clinic was open. Each day patients visited health facilities based on whether they had ILI symptoms or not and their risk level (low or high, Table 1). During the daytime, non-ILI patients only visited hospitals (not dedicated influenza clinics). ILI patients visited dedicated influenza clinics if they were open; otherwise, ILI patients visited hospitals and mixed with non-ILI patients. During the night, we assumed all clinics were closed; thus, all flu patients visited hospitals (e.g., EDs) if they needed care during this time. People seeking care at hospitals or clinics did not interact with their usual peer groups during the time of the healthcare consultation. Agents not hospitalized might have contact with their community group during day and night.

ILI patients may be hospitalized. Non-ILI patients remained in the hospital using length of stay (LOS) distributions: we assumed 92% stayed in the hospital for 6 hours on average, and 8% of them stayed for five days on average [29]. For non-hospitalized ILI patients, we assumed they were in hospital EDs and clinics for 6 hours and 3 hours on average, respectively. We also added a small random perturbation (less than 1.2 hours) to the average LOS for each patient to model the uncertainty. After the LOS, patients left the health facilities and returned to their routine contact network. See Tables 1 and 2 for the notations and parameters for baseline cases and sensitivity analysis.

For hospitalized patients, we considered two mixing modes at night with: (1) (MX) *mixing with family*, i.e., patients in the hospital interacted with their family members at night but with no other patients; (2) *mixing with patients*, i.e., ILI and non-ILI patients interacted in the same hospital without any household members.

Similar to other studies [19–22], we used calibration to estimate unknown transmission parameters, including coefficient of transmission, relative hazards of an infected agent in different disease stages, and social groups. Details are provided in the Appendix.

#### Settings of scenarios

We set the no-clinic scenario as the baseline and compared it to scenarios with clinics. The scenarios around clinics, length of time, and start week reflect operational decisions that may be impacted by the lead time necessary to organize resources. In this way, we can capture the lead time that may be associated with setting up dedicated clinics, and we capture the resources by measuring the total days of operation across multiple scenarios. The scenarios showing the effect of masks can be considered as a comparison intervention or related to hospital policies. Several other scenarios are used for sensitivity analysis, such as around the percentage of high-risk patients or reproductive rate.

We considered the temporal and spatial features of partially opening dedicated influenza clinics: initiation of operation date (*T_c_*), durations (*X_c_*), and locations (L_c_). In our model, clinics were categorized by location: clinics in metropolitan Atlanta (Cherokee, Clayton, Cobb, DeKalb, Douglas, Fayette, Fulton, Gwinnett, Henry, Rockdale counties) [34] as *Location Group I*, and clinics in other locations as *Location Group II*. The initiation of operation date and durations of clinic openings were predetermined in each experiment. Clinics within the same location group shared the same schedule. We calculated the clinic resource days by the number of counties open (10, 149, 159, or 0) times the number of weeks open (0, 4, 8, or 52) times five days per week for each scenario. The schedules of clinics are in Table 2.

While focusing on clinics, we also compared the effects of extensive use of masks by anyone in the hospitals or clinics. Two face masks are considered: surgical masks and N95. Compared to surgical masks, the N95 decreased susceptibility and infectivity nine times more strongly [31]. We assumed a certain percentage (*P_mk_*) of people in health facilities wore masks. See Table 2 for details.

Comparing scenarios, we determined the effects of opening clinics year-round (SCEN 1,29,31,33 versus SCEN 2,30,32,34), different clinic initiation dates and durations by location (SCEN 3-22), using masks in health facilities (SCEN 23-26,35-36), different mixing modes at night (SCEN 1,2,31,32 versus SCEN 29,30,33,34), and the proportion of high-risk people (SCEN 1,2,2930 versus SCEN 31,32,33,34),. Detailed descriptions of the scenarios are in Table 3. To minimize the stochastic effects during the initial phase of the outbreak, we seeded the model with 30 initial random cases. There were 30 replications for each scenario, where each replication simulated 365 days from 30 first infected cases randomly distributed in the network on day one.

**Table 3.**
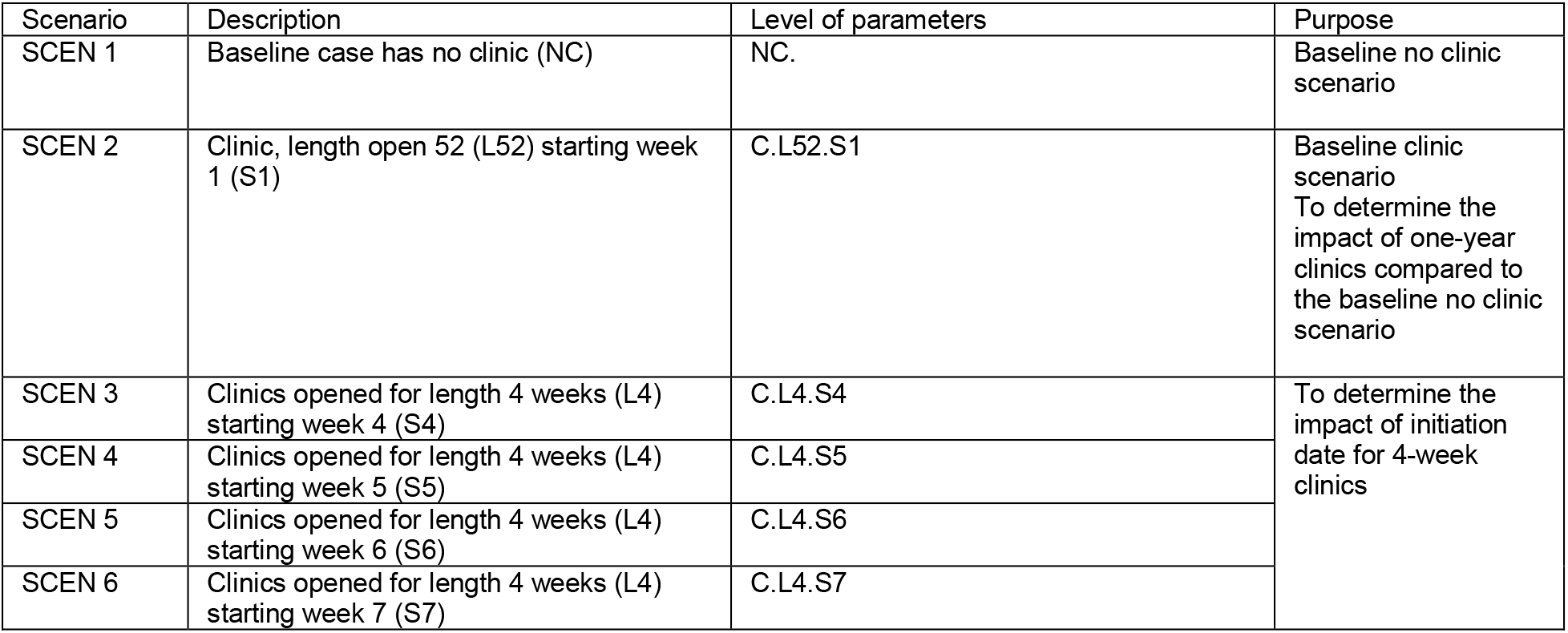
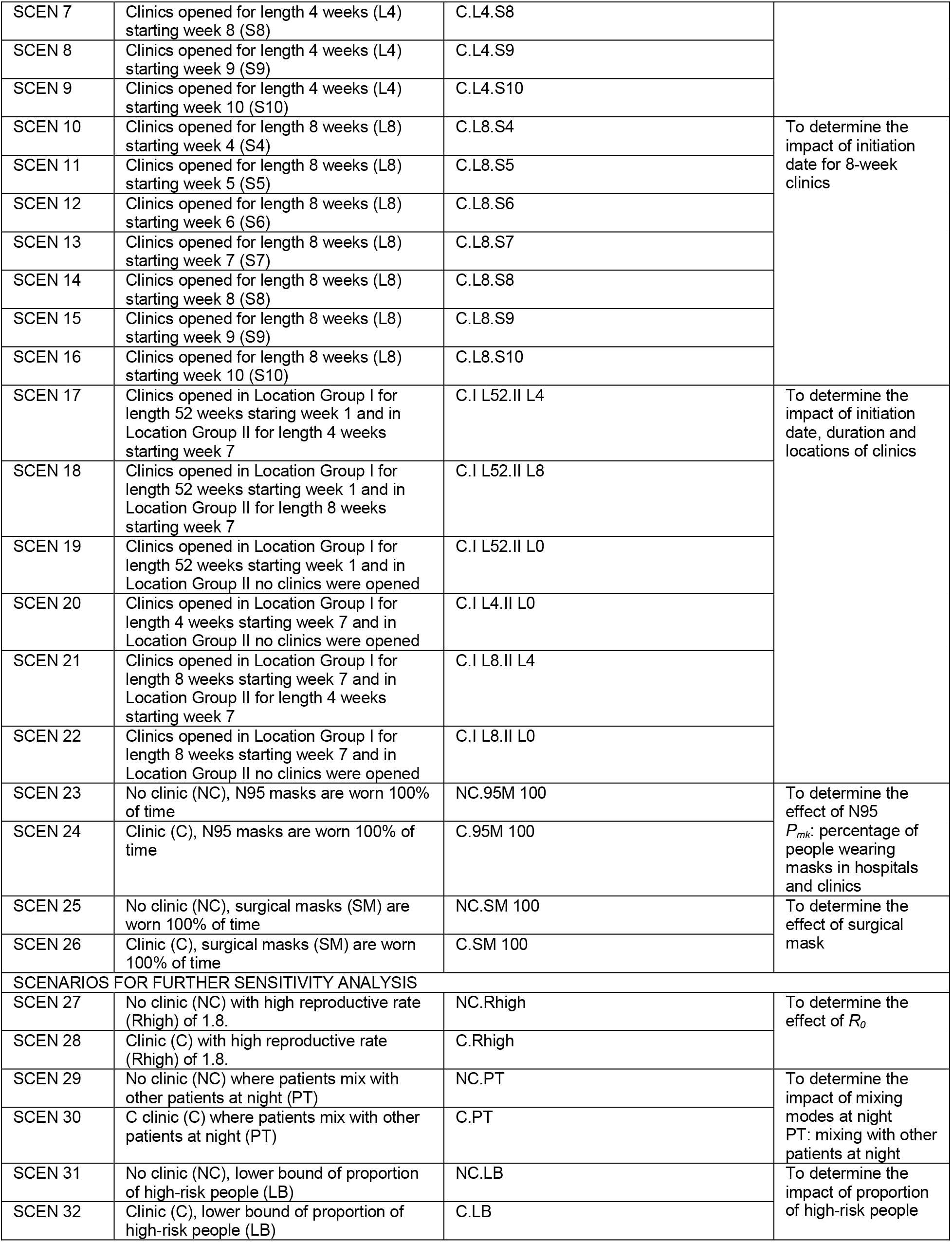
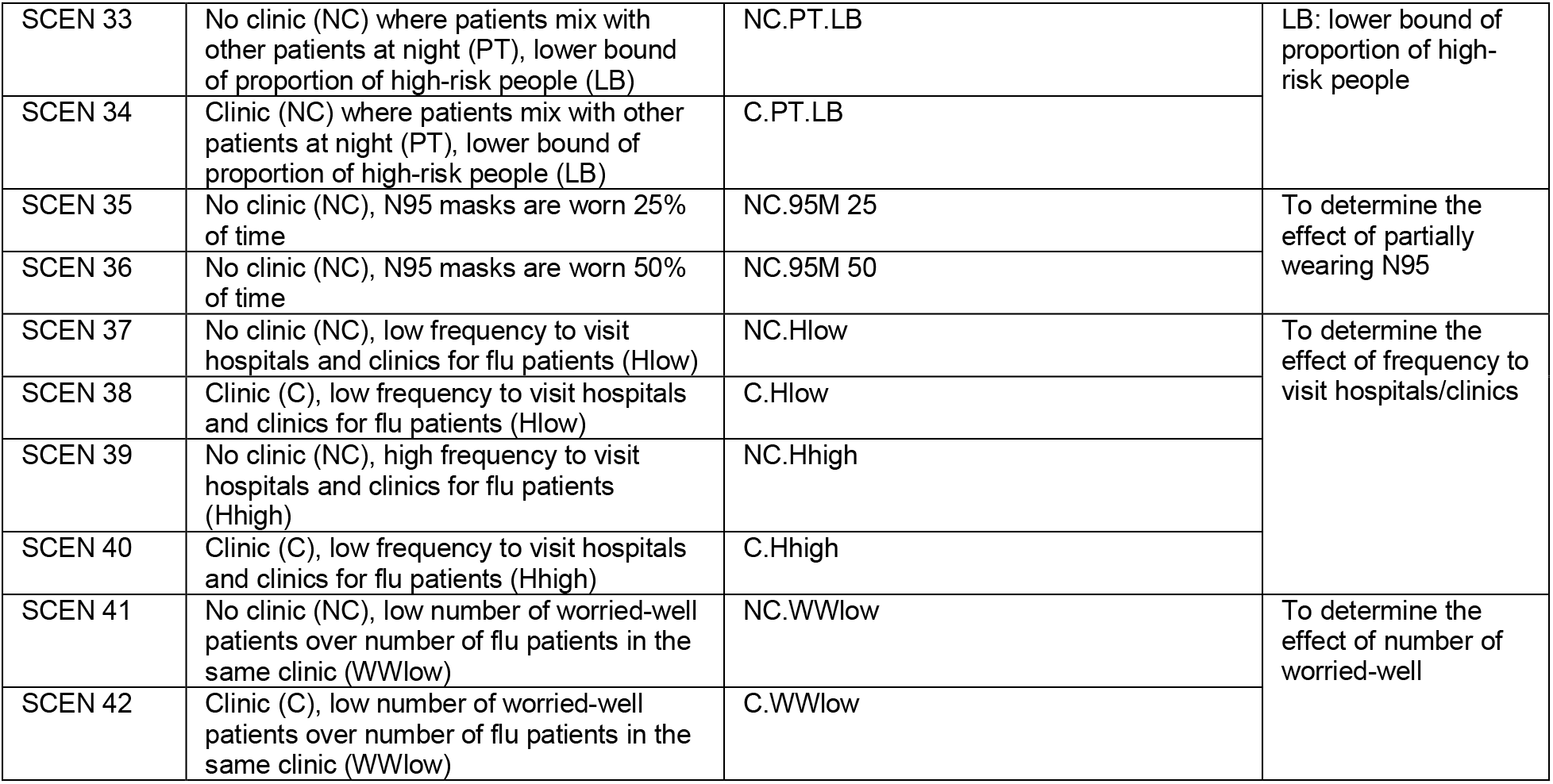
Settings of scenarios.

**Performance measures:** We compared the following criteria across scenarios:

- Peak prevalence of infected individuals (i.e., symptomatic and asymptomatic) and peak day (i.e., the first day of peak prevalence);
- Total attack rate (i.e., the cumulative percentage of individuals who have been infected by the virus);
- Total hospitalizations (i.e., the percentage of individuals who have ever been admitted for inpatient care);
- Total hospitalizations of children (i.e., the percentage of children who have been admitted for inpatient care);
- Infections in hospitals and clinics (i.e., the cumulative number of people who incurred infections in hospitals and clinics, including patients and companions).

For peak day, we used the median from the 30 replications as its estimator. For all other measures, we took the mean across 30 replications as the estimator. In addition, in the scenarios of timing and location, we reported the **relative benefit** achieved by partially opened clinics to that of one-year clinics as the ratio of the difference in performance measures of the relevant scenario (SCEN X) and SCEN 1 compared to that of SCEN 1 and 2, i.e., (SCEN1-SCENX)/(SCEN1-SCEN2). We used hospitalizations (which represent a severe outcome associated with influenza) as a proxy for mortality also.

**Sensitivity analysis**: We conducted a one-way sensitivity analysis using several parameters, including the frequency symptomatic flu patients visit health facilities (*q_f_*), the basic reproductive rate (*R*_0_) of influenza, and the proportion of worried-well people (*P_ww_*) with parameter values as presented in Table 2. The scenarios for sensitivity analysis (SCEN 29-42) are in Table 3.

We conducted two-sample paired t-tests in R (package version 3.2.2) to compare performance measures of pairs of scenarios and reported two-tailed p-values.

## Results

### Opening Clinics for One Year versus No Clinics (Table 4, Figs 1–2)

**Table 4.** Performance measures for baseline scenario and scenarios with or without clinics, different mixing modes at night, and proportion of high-risk people.

| Scenario | Level of Parameters | Peak prevalence (%) | Peak day | Total attack rate (%) | Hospitalization (%) | Hospitalization of children (%) | Infections in hospitals and clinics |
| --- | --- | --- | --- | --- | --- | --- | --- |
| SCEN 1 | NC | 3.41 | 65 | 55.82 | 3.07 | 5.21 | 330578 |
| SCEN 2 | C.L52.S1 | 3.19 | 68 | 54.64 | 3.01 | 5.08 | 281876 |

**Fig 1.**
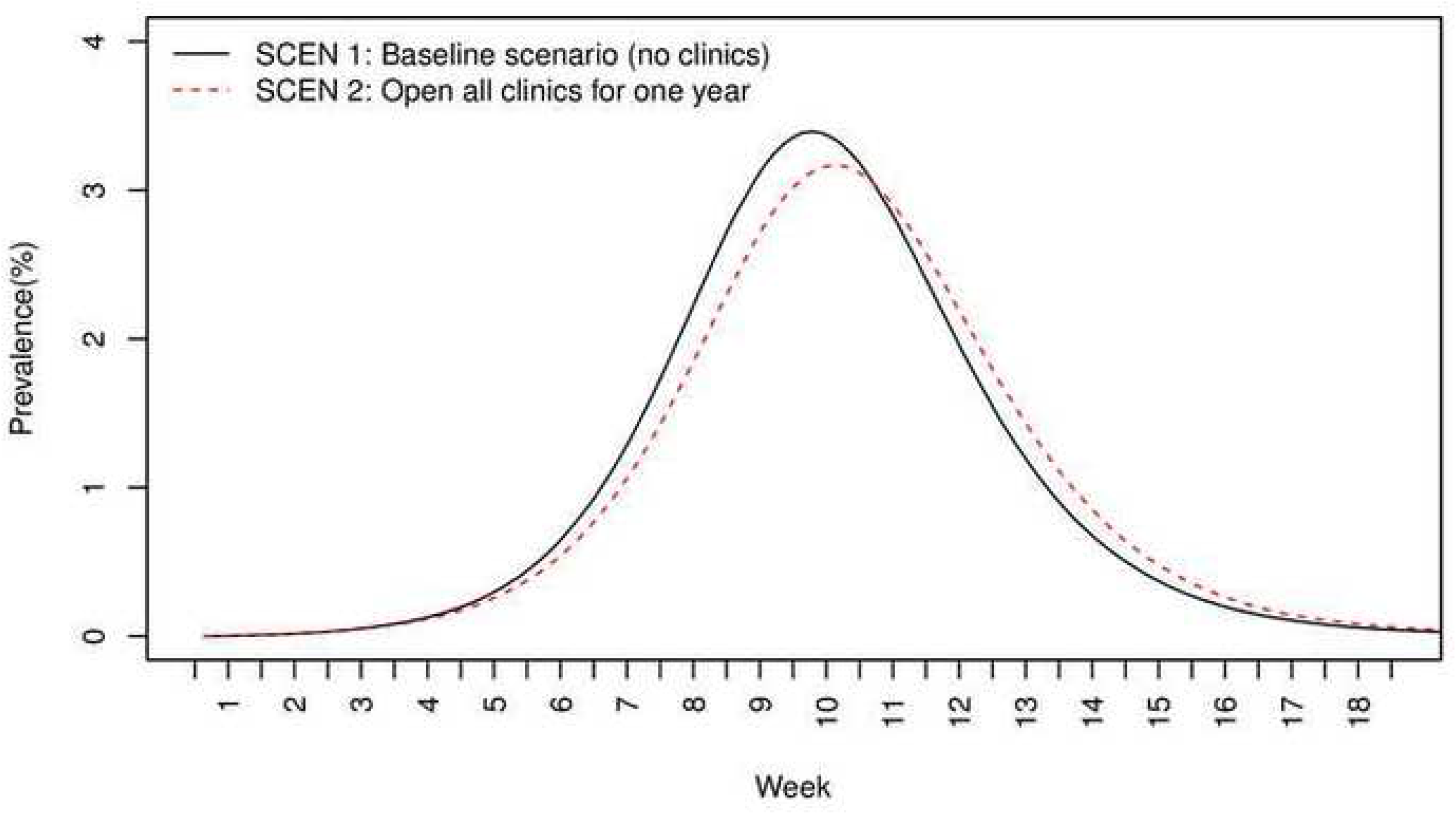
Prevalence (%) over time for SCEN 1 (no clinics) and SCEN 2 (one-year clinics).

**Fig 2.**
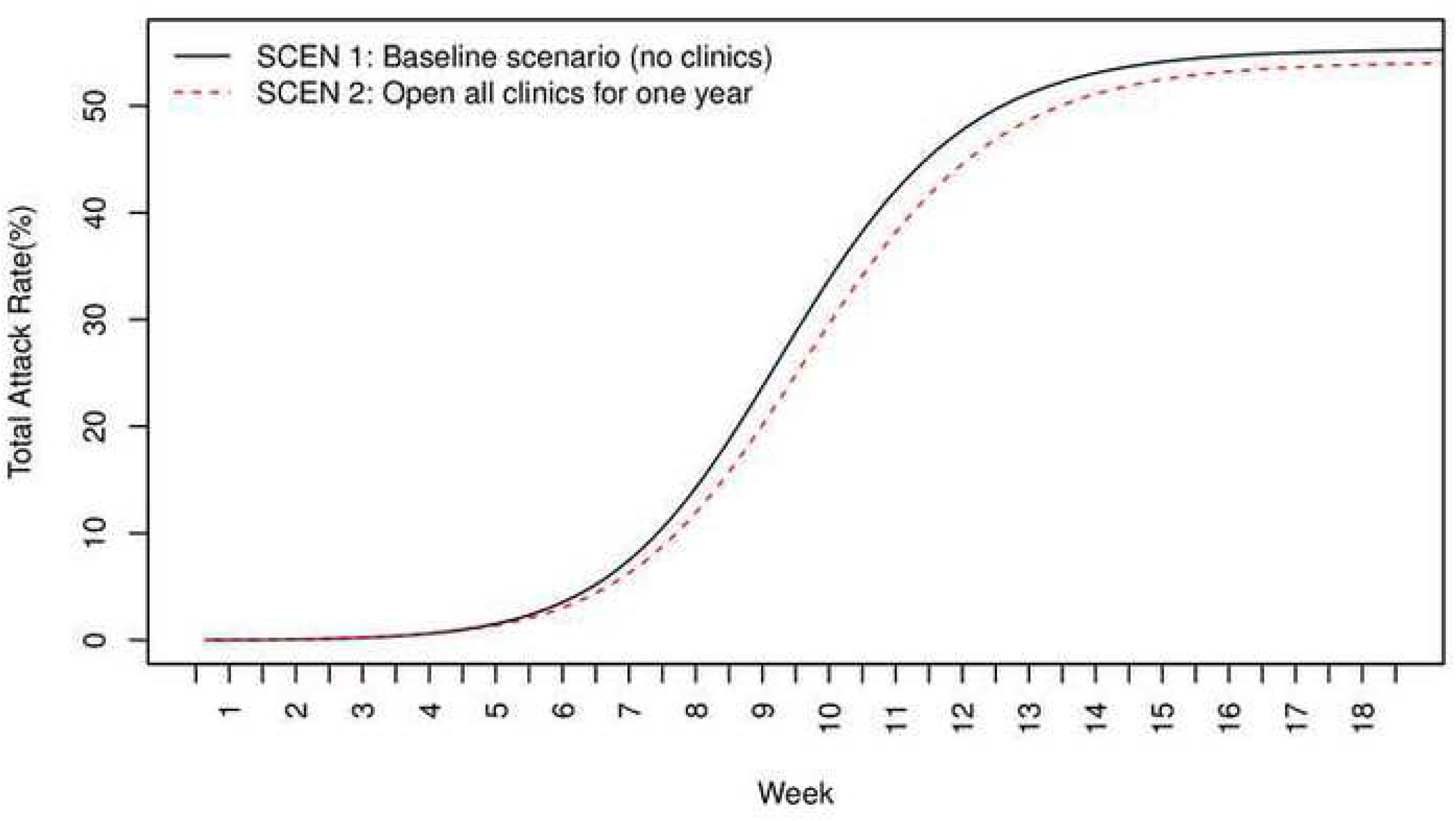
Total attack rates over time for SCEN 1 (no clinics) and SCEN 2 (one-year clinics).

Compared with no clinics (SCEN 1), opening all clinics for the entire simulation period of transmission (SCEN 2) had lower peak prevalence of infected individuals (3.19% with clinics versus 3.41% without, *p*<0.001), lower total attack rate (54.64% versus 55.82%, *p*<0.001), lower hospitalizations (3.01% versus 3.07% for the entire population, 5.08% versus 5.21% for children, *p*<0.001 respectively), and fewer infections occurring in hospitals and clinics (281876 versus 330578, *p*<0.001). Opening clinics also tended to delay the peak day of prevalence (68 versus 65, *p*<0.001). Figs 1–2 present the prevalence and total attack rate, respectively, from week 1 to week 18. Note that the peak days were in week 10 (SCEN 1-2, Fig 1).

### Benefits of Opening Clinics with Different Timing and Locations (Table 5, Figs 3–4)

The detailed results of opening clinics with different timing and locations are listed in Table 5. For 4-week scenarios, the best results occurred in SCEN 5 (open at week 6) with the lowest peak prevalence of 3.23%, SCEN 6 (open at week 7) with lowest infections in hospitals and clinics (306487), and SCEN 8 (open at week 9), which had the lowest total attack rate of 55.20%. For the 8-week scenarios, the best results were in SCEN 10 (open at week 4) with the lowest peak prevalence of 3.17%, SCEN 12 (open at week 6) with lowest infections in hospitals and clinics and low total attack rates (54.88%), and SCEN 13 (open at week 7) with the lowest total attack rates (54.86%) and low infections incurred in hospitals and clinics (294665).

**Table 5.** Performance measures for scenarios of clinics with different initiation of operation date, duration, or location along with baseline scenarios.

| Scenario | Level of Parameters | Clinic Days | Peak prevalence (%) | Peak day | Total attack rate (%) | Hospitalization (%) | Hospitalization of children (%) | Infections in hospitals and clinics |
| --- | --- | --- | --- | --- | --- | --- | --- | --- |
| SCEN 1 | NC | 0 | 3.41 | 65 | 55.82 | 3.07 | 5.21 | 330578 |
| SCEN 2 | C.L52.S1 | 41340 | 3.19 | 68 | 54.64 | 3.01 | 5.08 | 281876 |
| SCEN 3 | C.L4.S4 | 3180 | 3.31 | 67 | 55.63 | 3.08 | 5.20 | 319592 |
| SCEN 4 | C.L4.S5 | 3180 | 3.29 | 67 | 55.56 | 3.06 | 5.17 | 312562 |
| SCEN 5 | C.L4.S6 | 3180 | 3.23 | 67 | 55.41 | 3.05 | 5.17 | 308588 |
| SCEN 6 | C.L4.S7 | 3180 | 3.23 | 65 | 55.28 | 3.05 | 5.15 | 306487 |
| SCEN 7 | C.L4.S8 | 3180 | 3.28 | 65 | 55.24 | 3.05 | 5.15 | 308637 |
| SCEN 8 | C.L4.S9 | 3180 | 3.33 | 64 | 55.20 | 3.04 | 5.13 | 311290 |
| SCEN 9 | C.L4.S10 | 3180 | 3.40 | 64 | 55.32 | 3.05 | 5.16 | 315996 |
| SCEN 10 | C.L8.S4 | 6360 | 3.17 | 67 | 55.09 | 3.04 | 5.11 | 297205 |
| SCEN 11 | C.L8.S5 | 6360 | 3.20 | 66 | 54.98 | 3.03 | 5.12 | 293731 |
| SCEN 12 | C.L8.S6 | 6360 | 3.21 | 66 | 54.88 | 3.03 | 5.12 | 293154 |
| SCEN 13 | C.L8.S7 | 6360 | 3.23 | 65 | 54.86 | 3.02 | 5.10 | 294665 |
| SCEN 14 | C.L8.S8 | 6360 | 3.28 | 65 | 54.93 | 3.03 | 5.10 | 299343 |
| SCEN 15 | C.L8.S9 | 6360 | 3.33 | 64 | 54.97 | 3.03 | 5.12 | 305289 |
| SCEN 16 | C.L8.S10 | 6360 | 3.40 | 64 | 55.21 | 3.05 | 5.15 | 311689 |
| SCEN 17 | C.I L52.II L4 | 5580 | 3.22 | 66 | 54.96 | 3.02 | 5.11 | 296855 |
| SCEN 18 | C.I L52.II L8 | 8560 | 3.22 | 66 | 54.77 | 3.02 | 5.10 | 289756 |
| SCEN 19 | C.I L52.II L10 | 2600 | 3.33 | 66 | 55.42 | 3.06 | 5.17 | 311862 |
| SCEN 20 | C.I L4.II L0 | 200 | 3.33 | 65 | 55.60 | 3.06 | 5.16 | 319475 |
| SCEN 21 | C.I L8.II L4 | 3380 | 3.23 | 65 | 55.16 | 3.04 | 5.15 | 301911 |
| SCEN 22 | C.I L8.II L0 | 400 | 3.33 | 65 | 55.47 | 3.06 | 5.17 | 315450 |

**Fig 3.**
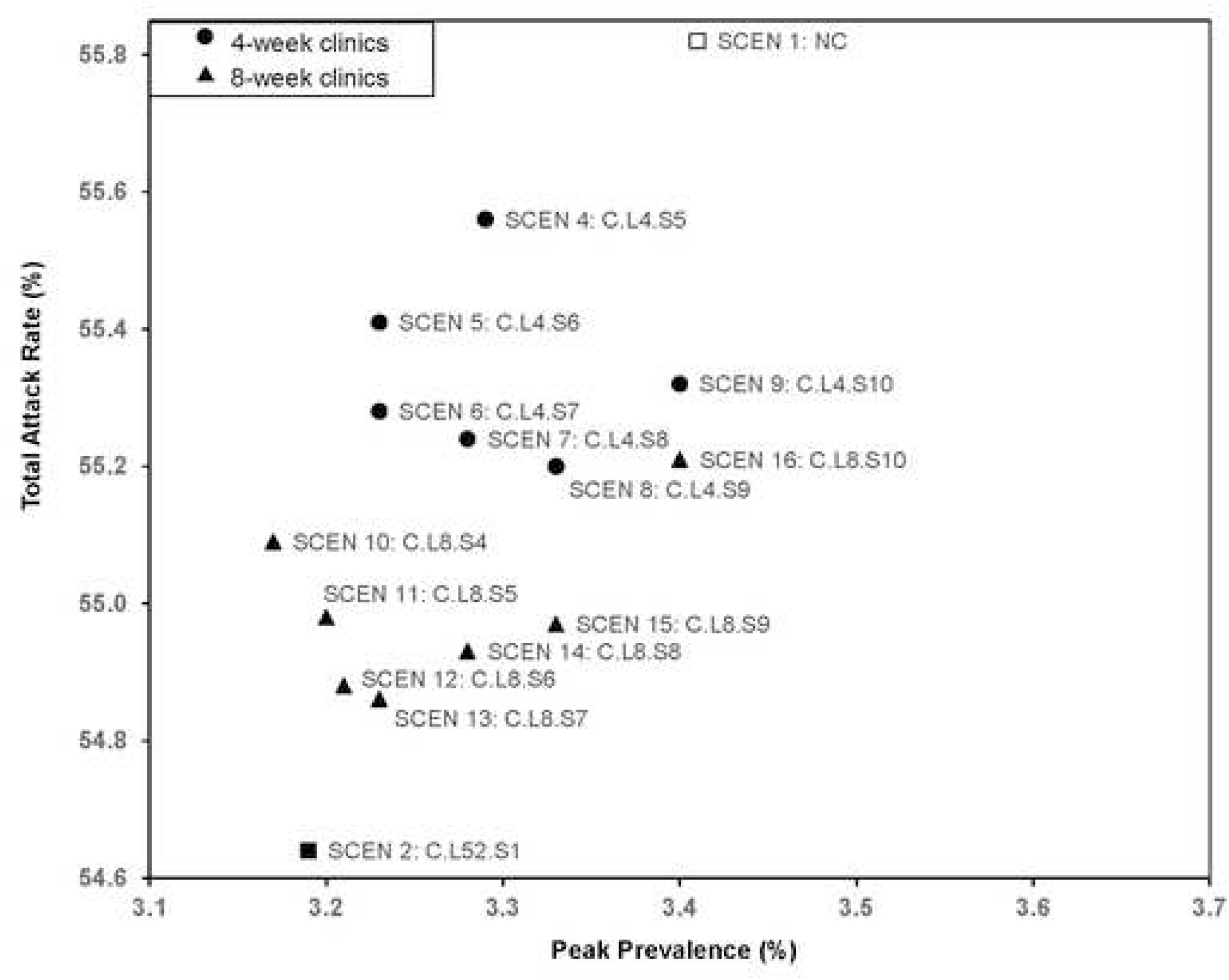
Total attack rate and peak prevalence of scenarios with 4(8)-week clinics open from weeks 4-10.

**Fig 4.**
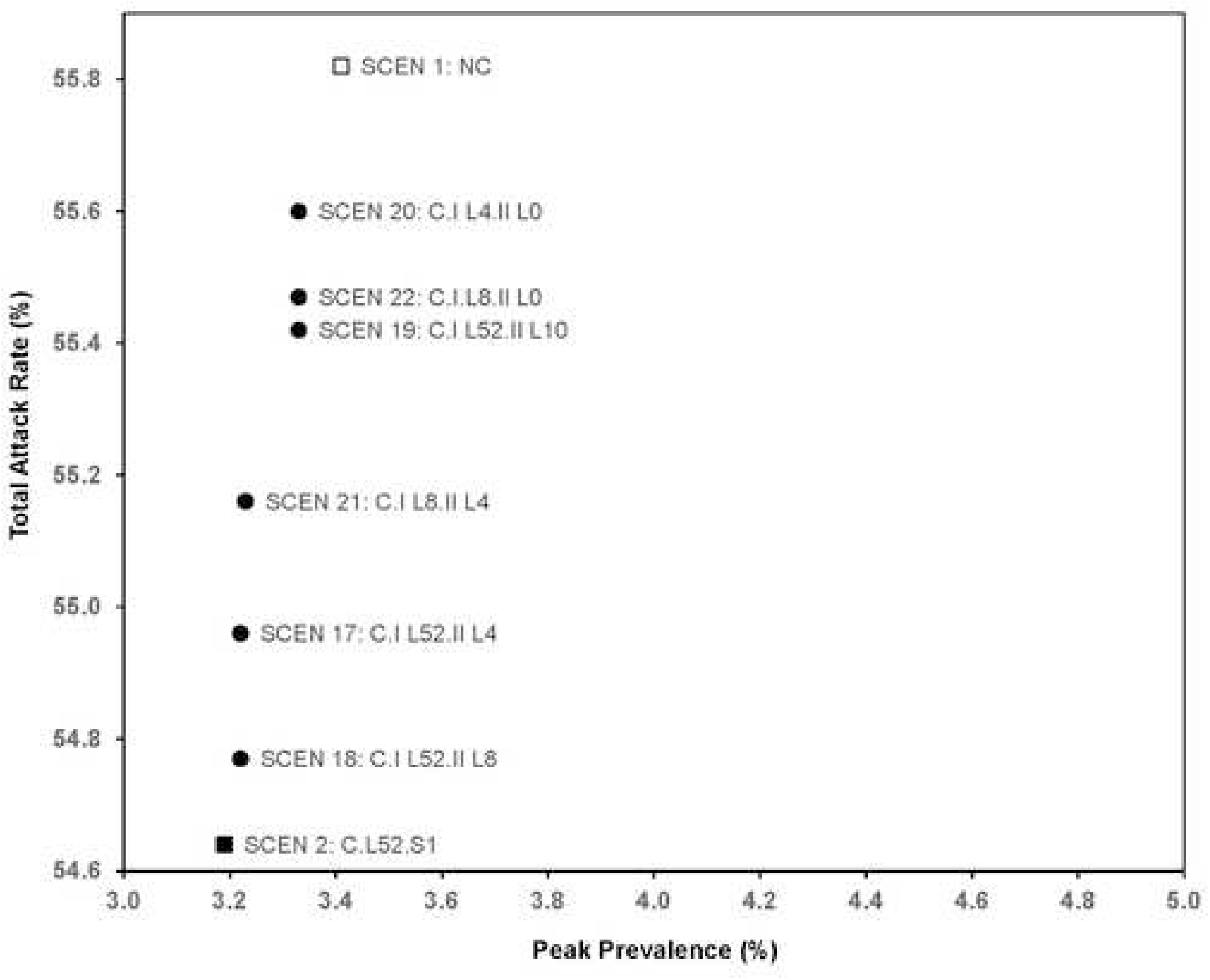
Total attack rate and peak prevalence of scenarios with location-based clinics.

The initiation of operation date of dedicated influenza clinics had some impact on performance measures. Comparing results of SCEN 3-16 (Fig 3), starting at week 7 (SCEN 6 for 4-week and SCEN 13 for 8-week clinics) had a low total attack rate (55.28% and 54.86%, respectively), low hospitalizations (3.05% and 3.02%, respectively), and low infections incurred in hospitals and clinics (306487 and 294665, respectively). In comparison, starting earlier (SCEN 3, open weeks 4-7, compared to SCEN 6 open weeks 7-10) could be worse, unless clinics are open longer (SCEN 10, open weeks 4-11). Overall, scenarios covering periods when prevalence was increasing and at its peak tended to be best (SCEN 5-8 versus SCEN 3,4,9, and SCEN 10-15 versus SCEN 16).

Regarding locations and durations (Fig 4), the total attack rate for opening clinics in the metropolitan Atlanta region only for one year (SCEN 19) was 55.42%, which had a relative benefit of 34% of the reduction in total attack rate by opening clinics for one year in the entire state (SCEN 2). The results of SCEN 18, where clinics in the metropolitan Atlanta region were open for one year and in other locations for eight weeks starting at week 7, gained more of the relative benefits of opening clinics everywhere for a year, specifically 89%.

### Effects of Masks Compared to Opening One-Year Clinics (Table 6, Fig 5)

Opening one-year clinics (SCEN 2) had a stronger effect on total attack rate (54.64%) than fully wearing surgical masks (SCEN 25, *p*<0.001) and 25% wearing N95 (SCEN 35, *p*<0.001), while 50% wearing N95 (SCEN 36, *p*<0.001) was slightly better than fully opening clinics (SCEN 2). Wearing masks tended to reduce infections incurred in hospitals and clinics as well as hospitalizations. Fully wearing N95 (SCEN 23) dominated opening clinics for a full year (SCEN 2) in both peak prevalence and total attack rate (*p*<0.001), although the combined effect of masks and clinic could be greater (SCEN 24). Table 6 summarizes the effects of masks.

**Table 6.** Performance measures for scenarios with masks.

| Scenarios | Level of Parameters | Peak prevalence (%) | Peak day | Total attack rate (%) | Hospitalization (%) | Hospitalization of children (%) | Infections in hospitals and clinics |
| --- | --- | --- | --- | --- | --- | --- | --- |
| SCEN 23 | NC.95M 100 | 3.01 | 68 | 52.97 | 2.90 | 4.90 | 169843 |
| SCEN 24 | C.95M 100 | 2.94 | 70 | 52.53 | 2.88 | 4.88 | 149232 |
| SCEN 25 | NC.SM 100 | 3.39 | 65 | 55.61 | 3.06 | 5.17 | 313313 |
| SCEN 26 | C.SM 100 | 3.18 | 68 | 54.47 | 3.00 | 5.07 | 268401 |
| SCEN 35 | NC.95M 25 | 3.31 | 66 | 55.07 | 3.04 | 5.14 | 287308 |
| SCEN 36 | NC.95M 50 | 3.19 | 67 | 54.35 | 2.99 | 5.05 | 246415 |

**Fig 5.**
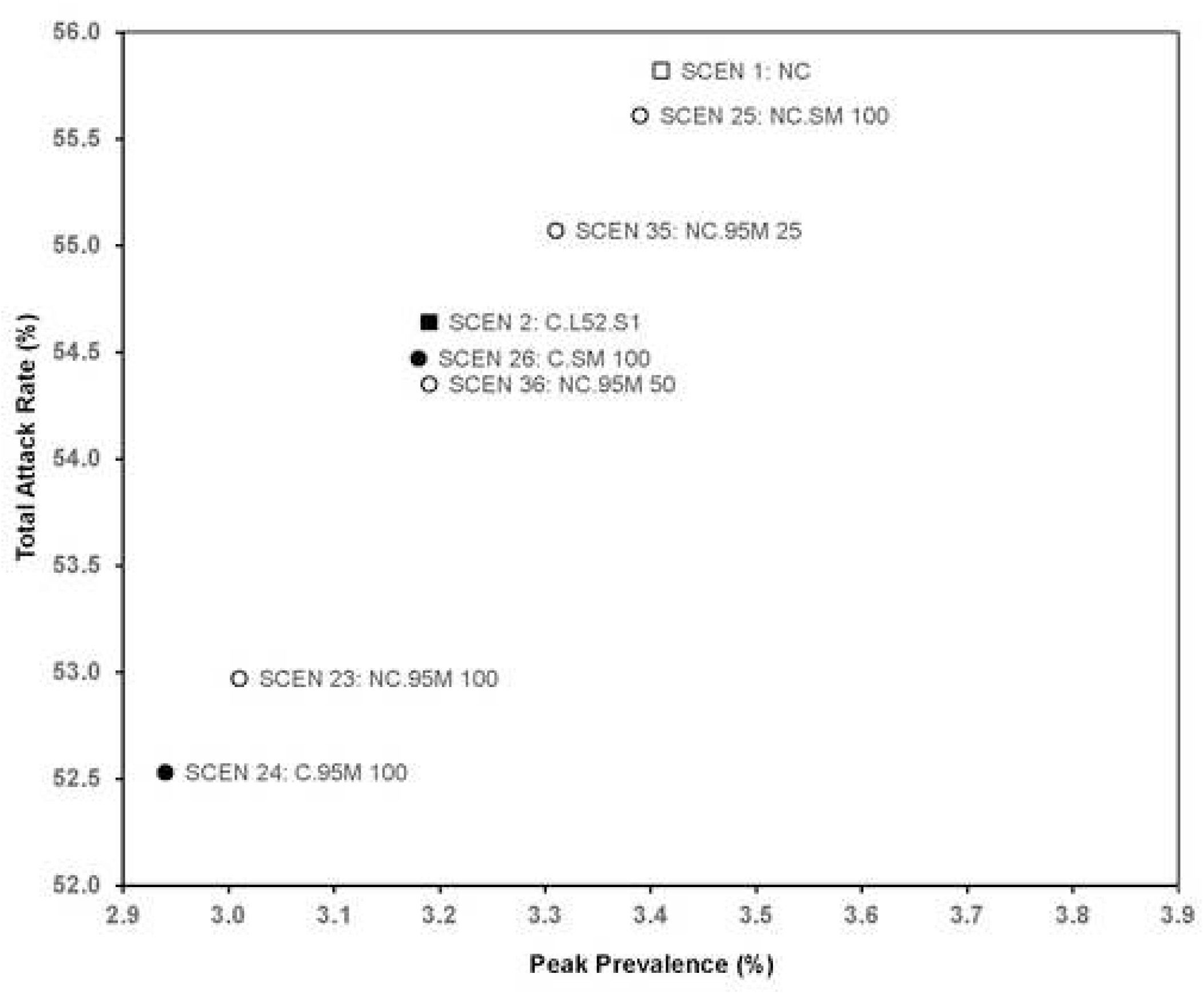
Total attack rate and peak prevalence of scenarios with masks.

### Sensitivity Analysis (Table 7, Figs 6–8)

For a higher reproduction rate (*R*_0_ = 1.8), the total attack rate increased from the base case (SCEN 1 with 55.82%) to 71.15% for no clinics (SCEN 27) or to 70.44% with 1-year clinics (SCEN 28). When patients mixed with family members at night, by opening clinics for one year, the total attack rate reduced 1.18% (SCEN 1 versus SCEN 2, *p*<0.001). When patients mixed with other patients at night, opening clinics reduced the total attack rate by 0.40% (SCEN 29 versus SCEN 30, *p*<0.001). Similarly for using the lower bound on high-risk patients, having clinics open for a year reduced the total attack rate by 1.00% when patients mixed with family members at night (SCEN 31 versus SCEN 32, *p*<0.001), and by 0.50% when patients mixed with other patients at night (SCEN 33 versus SCEN 34, *p*<0.001). The total attack rate and peak prevalence of the SCEN 1-2 and 29-34 are presented in Fig 7.

The total attack rates for low, medium, and high chance of visiting hospitals and clinics with 1-year clinics (SCEN 38, 2, and 40) were 55.55%, 54.64%, and 54.21%. If clinics brought in fewer worried-well (SCEN 42), the total attack rate was 55.20%, a little higher (*p*<0.001) than that for SCEN 2, which had more worried-well at *R*_0_ = 1.5. Table 7 summarizes the effects of clinics under various rates of visiting hospitals and clinics, a higher reproduction rate, or a lower proportion of worried-well.

**Table 7.** Sensitivity analysis.

| Scenarios | Level of Parameters | Peak prevalence (%) | Peak day | Total attack rate (%) | Hospitalization (%) | Hospitalization of children (%) | Infections in hospitals and clinics |
| --- | --- | --- | --- | --- | --- | --- | --- |
| SCEN 27 | NC.Rhigh | 6.66 | 48 | 71.15 | 3.94 | 6.59 | 333422 |
| SCEN 28 | C.Rhigh | 6.36 | 50 | 70.44 | 3.88 | 6.49 | 297954 |
| SCEN 29 | NC.PT | 2.90 | 69 | 51.87 | 2.84 | 4.76 | 138049 |
| SCEN 30 | C.PT | 2.83 | 71 | 51.47 | 2.82 | 4.76 | 104961 |
| SCEN 31 | NC.LB | 3.37 | 66 | 55.40 | 2.93 | 4.74 | 292131 |
| SCEN 32 | C.LB | 3.16 | 68 | 54.40 | 2.88 | 4.64 | 254583 |
| SCEN 33 | NC.PT.LB | 2.91 | 70 | 51.91 | 2.73 | 4.39 | 118929 |
| SCEN 34 | C.PT.LB | 2.84 | 70 | 51.41 | 2.71 | 4.37 | 93930 |
| SCEN 37 | NC.Hlow | 3.42 | 66 | 56.01 | 3.09 | 5.21 | 309215 |
| SCEN 38 | C.Hlow | 3.33 | 67 | 55.55 | 3.06 | 5.16 | 284940 |
| SCEN 39 | NC.Hhigh | 3.43 | 65 | 55.72 | 3.07 | 5.19 | 342944 |
| SCEN 40 | C.Hhigh | 3.10 | 69 | 54.21 | 2.98 | 5.04 | 281840 |
| SCEN 41 | NC.WWlow | 3.41 | 65 | 55.82 | 3.07 | 5.21 | 330578 |
| SCEN 42 | C.WWlow | 3.26 | 67 | 55.20 | 3.03 | 5.11 | 284555 |

**Fig 6.**
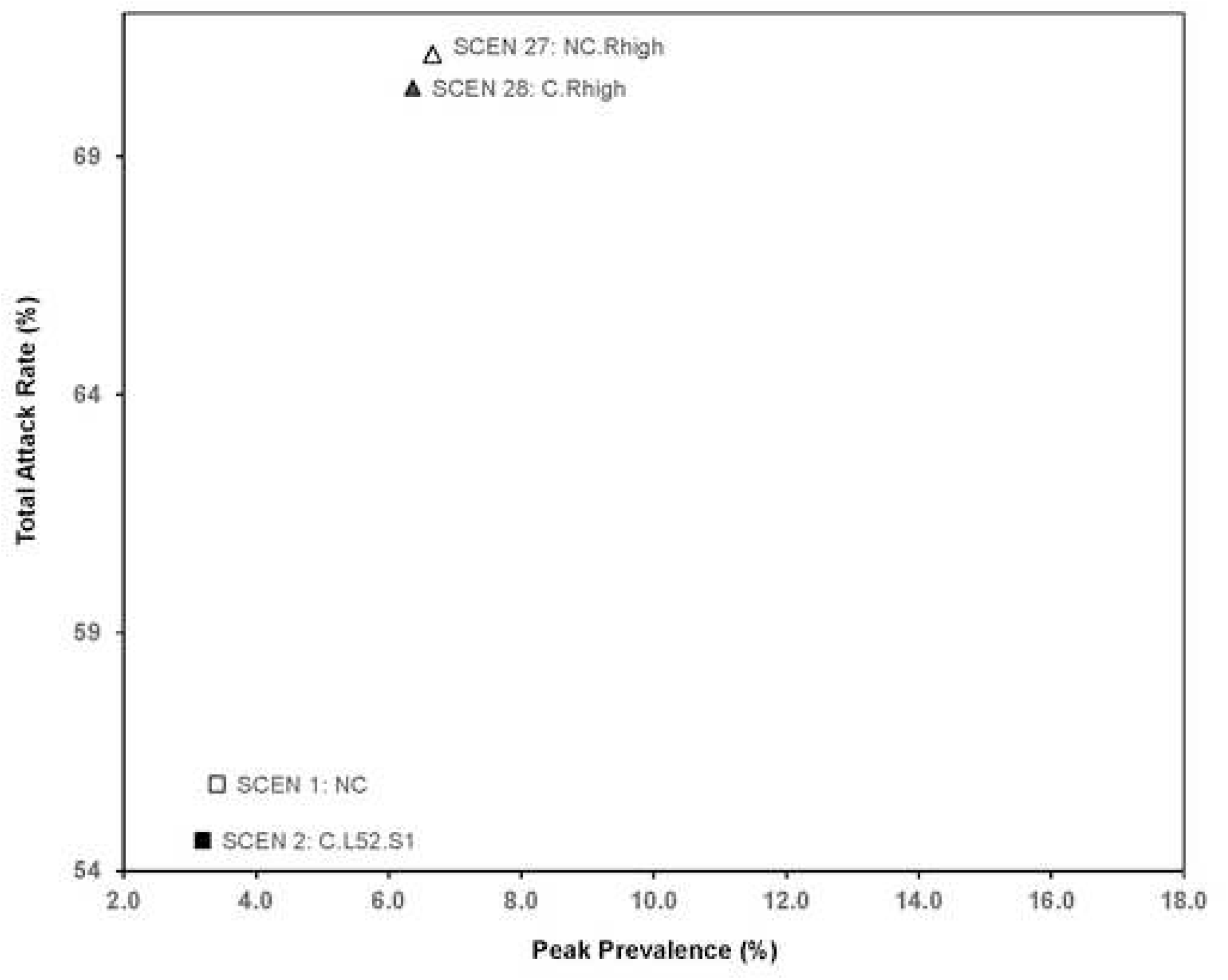
Total attack rate and peak prevalence of scenarios for sensitivity analysis (higher reproductive rate).

**Fig 7.**
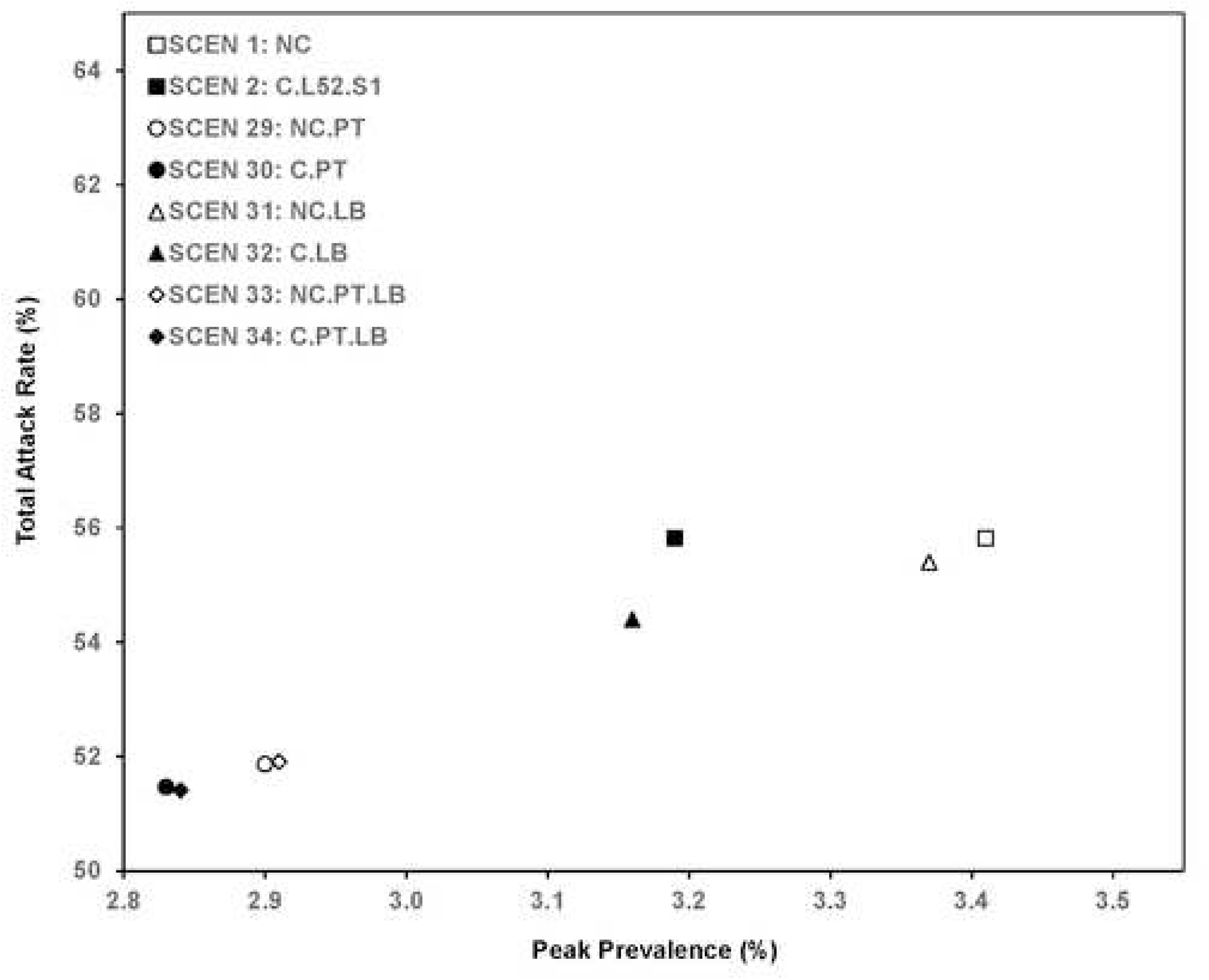
Total attack rate and peak prevalence of scenarios without clinics and with 1-year clinics.

**Fig 8.**
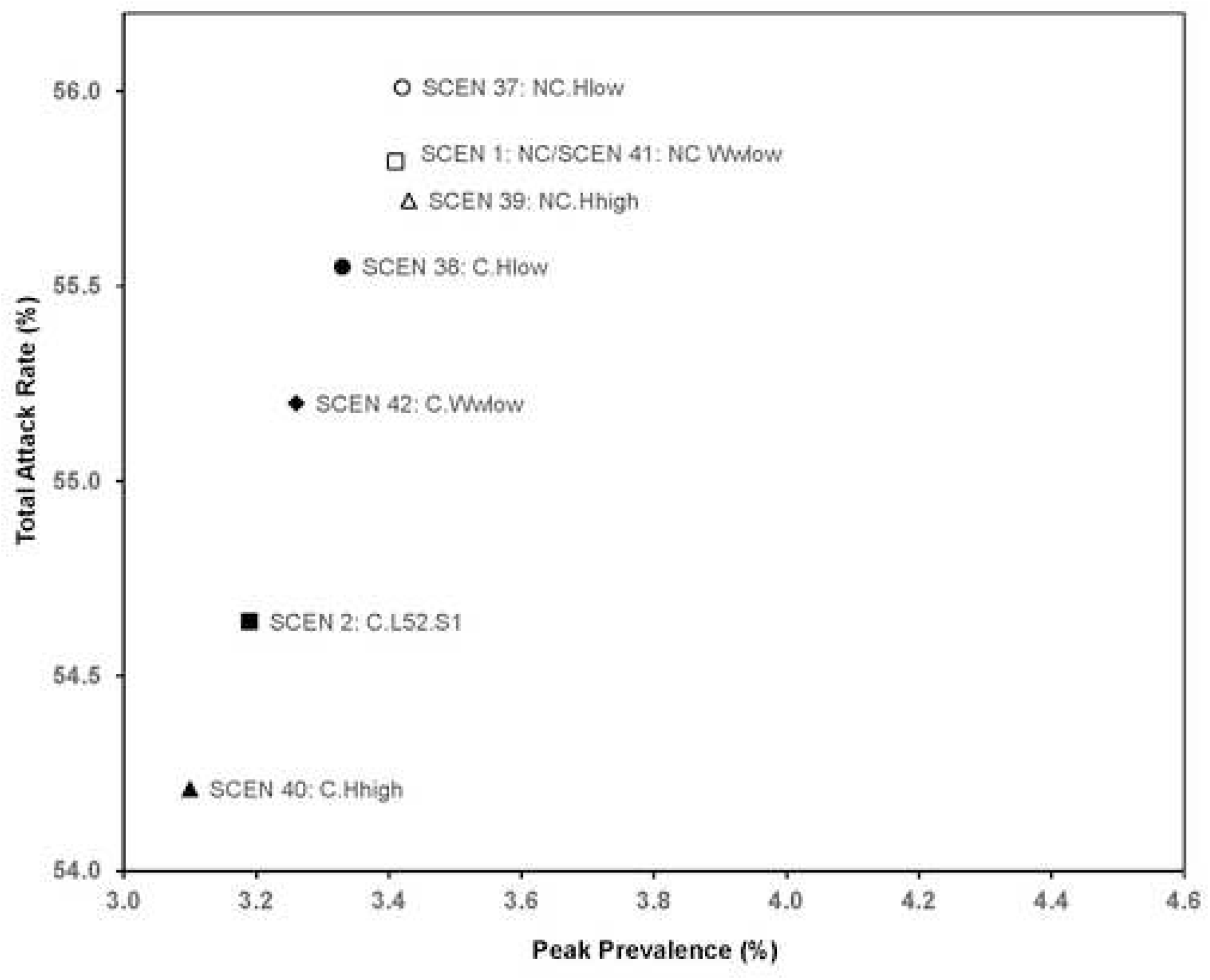
Total attack rate and peak prevalence of scenarios for low/high visit frequency, lower worried-well proportion.

Table 8 gives values for paired t-tests on multiple performance measures for many paired scenarios. We found statistically significant differences in the Total Attack Rate for all scenarios with a dedicated clinic as compared to a similar scenario without, at the 5% level or stronger. For a state that has a population of approximately 10 million, the difference in the baseline clinic case would be about 100,000 cases averted using 41,340 clinic days. For hospitalizations, we also found statistically significant differences for all scenarios with clinics except for SCEN 36, which was inconclusive. For a population of 10 million, the baseline case with clinics open would translate to about 6,000 hospitalizations averted. Comparing scenarios where clinics were open for a short time (4 weeks, SCEN 3) to a longer time (8 weeks, SCEN 10), the difference in total attack rate and hospitalizations would translate to about 50,000 cases averted and 4,000 hospitalizations averted, using 3,180 and 6,360 clinic days. The scenario with full clinics is better than that with clinics only in location set I (SCEN 19), translating into approximately 78,000 additional cases (or 5,000 hospitalizations) averted. Having full dedicated clinics was better than having 100% surgical masks (SCEN 25) with 97,000 cases and 5,000 hospitalizations averted, but having 100% N95 masks was better than having dedicated flu clinics, translating to approximately 167,000 cases and 11,000 hospitalizations averted.

**Table 8.**
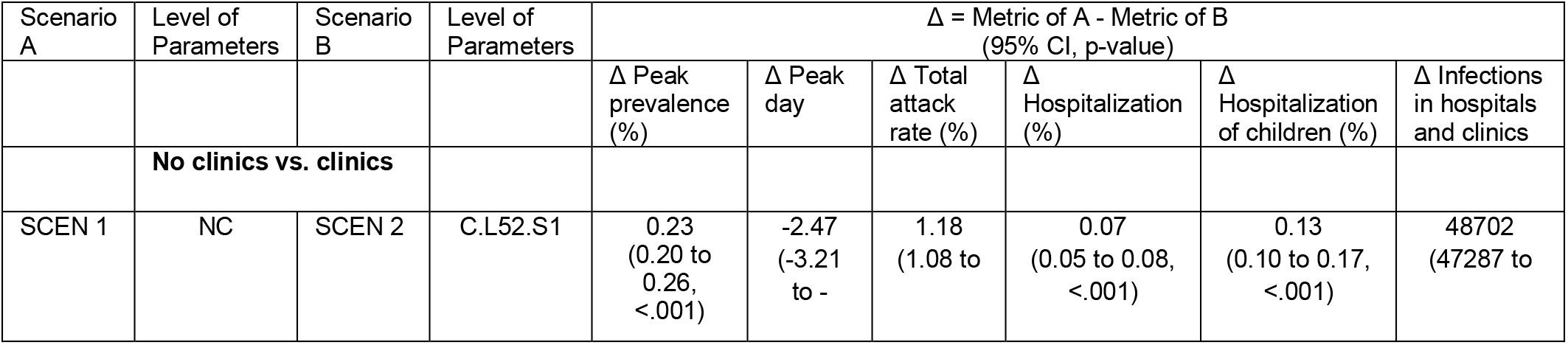
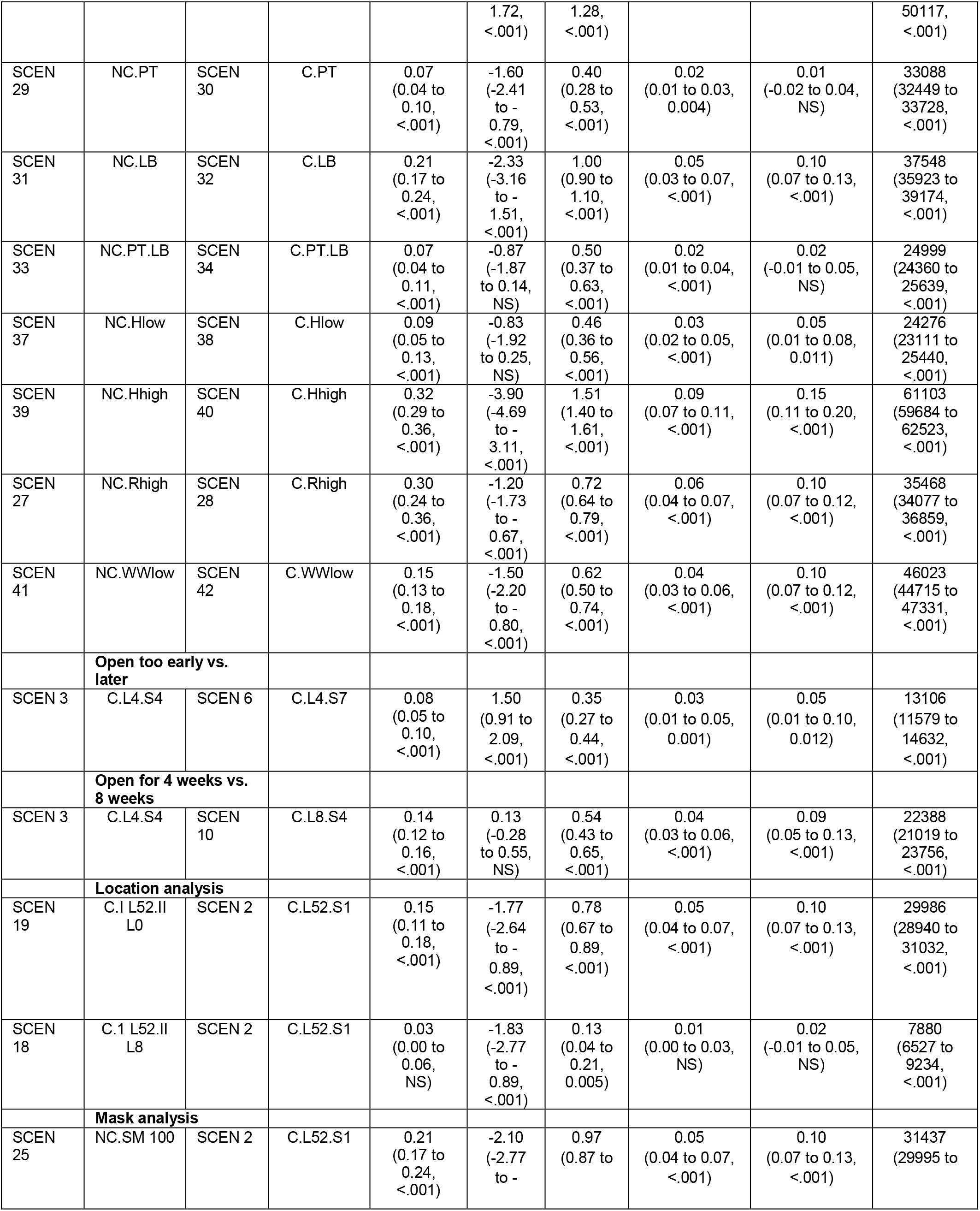
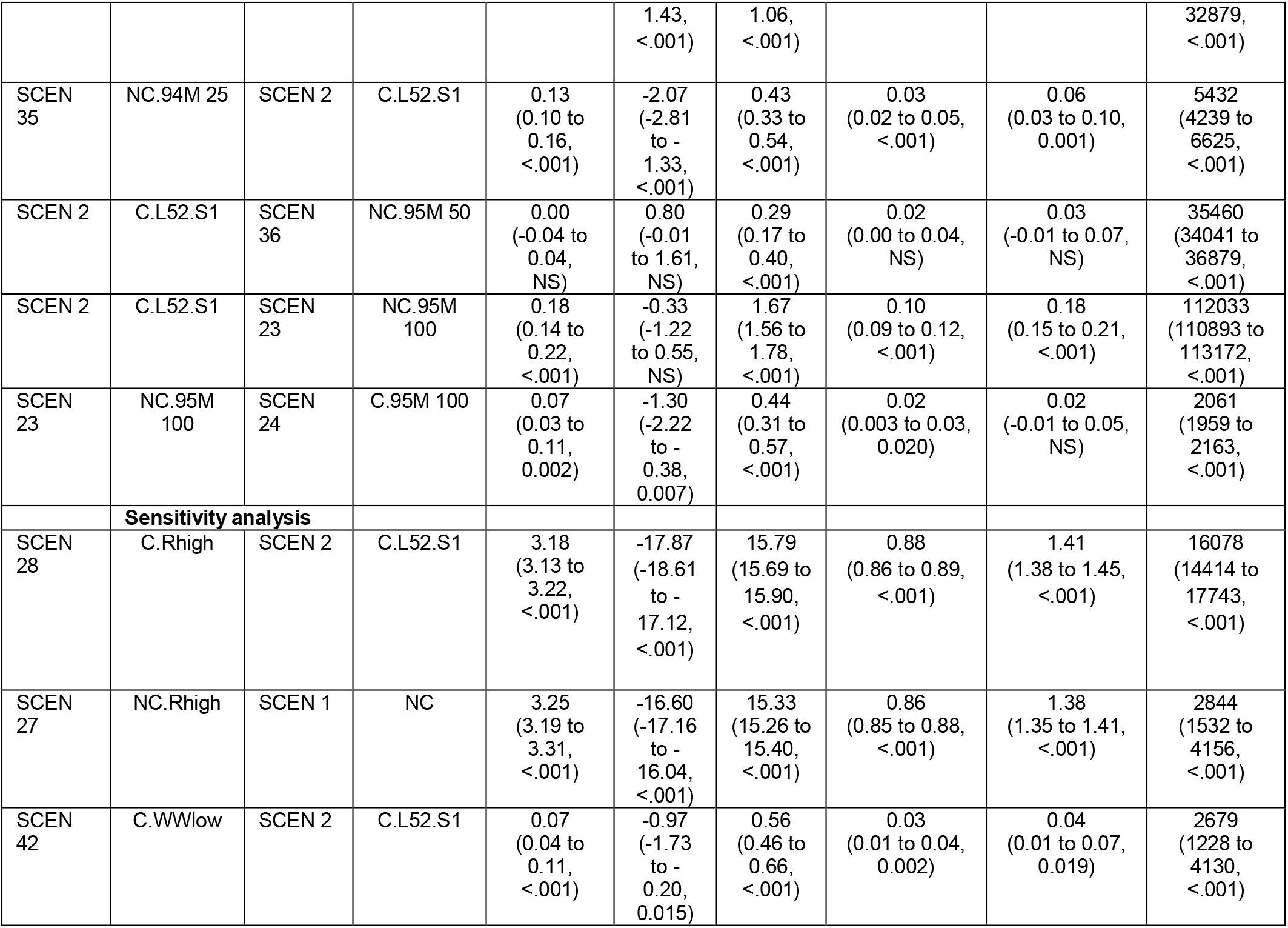
Paired t-tests, two-tailed (The alternative hypothesis is that the performance measures of Scenarios A and B are different; for measures other than peak day having a smaller performance measure is better. NS (not significant) denotes that the p-value>0.05).

## Discussion

The main goal of this research was to guide stakeholders on resource allocation by determining the impact of dedicated influenza clinics on the spread of disease during a pandemic under different scenarios. Dedicated clinics may also offer some additional benefits. For example, they could be used as a distribution point for medical countermeasures such as anti-virals. Additionally, such clinics could help with the availability of masks, which could be prioritized for personnel working in clinics dedicated to infectious disease. Clinics have the potential to draw the worried-well but also isolate flu patients from non-ILI patients. The results of comparing SCEN 1,29,31,33 (no clinic scenarios) versus 2, 30,32,33 (clinics open for one year) give an unequivocal conclusion that opening clinics for the duration of the epidemic can significantly reduce peak prevalence, total attack rate, hospitalizations, and the number of infections in healthcare facilities. The total attack rate is the lowest for scenarios that open clinics for the longest time period including SCEN 2, followed by SCEN 18, 12, and 13. The peak prevalence tends to be lowest for scenarios with clinics open the entire time period (SCEN 2).

The peak can also be affected by clinics with specific opening times (e.g., SCEN 10), which is similar to putting an intervention in place for a limited time. The total attack rate is lowest when clinics are open a long time or cover the majority of the peak (e.g. SCEN 2 or SCEN 11-15).

While changing the attack rate from 55.82% to 51.41% may seem like a small change, for a population in one state the size of Georgia, this would translate to approximately 500,000 cases averted. Similarly, the change in hospitalization and mortality means that the best case intervention reduces hospitalization from 330,578 to 93,930 for a state the size of Georgia. The impact from dedicated clinics is not as much as a voluntary 8-week quarantine [see Appendix of reference 5] or vaccine that covered 20% or more of the population [35]. Yet averting more than 100,000 cases and thousands of hospitalizations may still be needed. In addition, having clinics open can delay the peak day of disease spread, which provides more time for the preparation of resources. These conclusions also hold for different visit rates to hospitals (SCEN 37,1,39 versus 38,2,40), fewer worried-well (SCEN 41 versus 42), and higher *R*_0_ (SCEN 27 versus 28).

In practice, with limited resources such as healthcare personnel, the operation of dedicated influenza clinics during a pandemic should maximize resource utilization. In the presence of dedicated clinics, there may be additional ways to serve the worried-well, such as encouraging them to call their practitioner for advice. Another advantage of the dedicated clinics is that they could free up resources for non-influenza related healthcare needs. We studied the effect of clinics opened after a period of time to examine the role that lead time has on outcomes. An alternative would be opening clinics after the epidemic passed some threshold of cases, where the threshold may occur sooner for high values of R_0_. This requires good knowledge of the R_0_ and the true cases in the population. Opening for one year may not be practical or needed.

Rather, most of the effects of clinics can be achieved by carefully selecting start time and duration based on the pandemic dynamics. In particular, a goal should be to cover the periods when prevalence is increasing and at its peak. While in real-time, the peak is difficult to know, the usual rule of starting early enough (e.g., week 7) applies, and covering a number of weeks of high prevalence offers significant benefits even with limited resources (SCEN 13 vs. 2). Concentrating some clinical resources everywhere with additional resources in heavily populated areas is also a good strategy to consider (SCEN 18 vs. 2). If it is possible to reduce the lead time to set up clinics or reduce the resources associated with running a clinic, then additional benefits may be achieved, as demonstrated by the timing analysis.

Mixing patterns in hospitals and clinics impact disease control (Fig 7). If hospitalized flu-infected patients have contact with their uninfected family members at night (SCEN 1,2,31,32), transmission to household contacts and subsequently to members of peer groups of infected family members results in propagation of infection. Conversely, if flu-infected patients mix only with other flu-infected patients at night (SCEN 29,30,33,34), the hospitals can have a strong isolation effect, which can reduce disease spread in the population. Clinics may bring in worried-wells but may not necessarily increase infection transmission. The worried-well may affect disease spread in two ways. First, they are removed from their peer groups while they visit a healthcare facility and cannot get infected by their peer group. However, they have an elevated risk of being exposed to infections within clinics. When more worried-well enter clinics, the clinics create isolation for the worried-well away from their peer groups. Our model shows that increasing the number of worried-well (*P_ww_* from 0.2 to 0.5, SCEN 42 versus SCEN 2) decreases the total attack rate (55.20% to 54.64%, *p*<0.001, Table 8, Fig 8), which implies that the reduction in infection by removal from peer groups is larger than the increased risk of infection by being present in a clinic. We find a reduction in hospitalizations, which is also a proxy for potential changes in mortality.

The results on masks are useful to consider. Having 50% of people in health facilities wearing N95 masks gives a similar reduction in attack rate as opening clinics for a full year (SCEN 36 and SCEN 2) while wearing 100% surgical masks (SCEN 25) is a little less effective than opening clinics fully (SCEN 2). Moreover, the resources required for masks would likely be much less than clinics, as long as a sufficient supply of N95 masks is stockpiled or available and if patients would wear masks according to the guidelines within healthcare facilities.

## Limitations

Modeling brings the usual limitations, including that it is based on assumptions such as mixing patterns and transmission rates. Our specific study also assumes people go to the regional hospital that is geographically close. We assume each hospital sets up one dedicated influenza clinic that has the resources to serve everyone. We validated our model against pandemics of previous decades but acknowledge that travel patterns and contact may have been different during that time. For some parameters, we do not have an accurate estimate, so we test different values to examine the robustness of our results. However, we could not test all combinations because of the large number of scenarios.

## Conclusions

Public health benefit results from the opening and operating of dedicated influenza clinics to diagnose and manage people with influenza-like illness during an influenza pandemic. Transmission of pandemic influenza infections is reduced when dedicated clinics are fully operational during the times of the greatest prevalence of the pandemic. Alternative strategies include operating clinics during the periods of highest prevalence during the pandemic, operating clinics based on population density, and wearing N95 in healthcare facilities.

## Data Availability

All data are fully available without restriction

## Acknowledgments

We thank Ali Ekici and Joseph T. Wu for sharing their C++ code. We extend their code of a basic disease spread model to study the impacts of opening dedicated influenza clinics.

## Author contributions

Pengyi Shi created the initial modifications to the simulation to allow dedicated influenza clinics. Jia Yan carried out all analyses in the paper, drafted the manuscript, critically reviewed and revised the manuscript, and approved the final manuscript as submitted.

Andi L. Shane provided empirical data and expertise related to hospital operation and clinics, critically reviewed and revised the manuscript, and approved the final manuscript as submitted. Pinar Keskinocak and Julie Swann supervised all analyses and preparing of the manuscript, critically reviewed and revised the manuscript, and approved the final manuscript as submitted.

## Disclaimers

No conflict of interests

## Funding information

This research has been supported in part by a seed grant from Georgia Tech and by the following Georgia Tech benefactors: William W. George, Andrea Laliberte, Claudia L. and J. Paul Raines, and Richard E. “Rick” and Charlene Zalesky. This research has also been supported by Edward P. Fitts and the A. Doug Allison Distinguished Professorship. Any opinions, findings, conclusions, or recommendations are those of the authors and do not necessarily reflect the views of the funding organizations.

## Supporting information

**S1 File. Appendix of the Impact of Opening Dedicated Clinics on Disease Transmission during an Influenza Pandemic**.

## S1. Appendix of the Impact of Opening Dedicated Clinics on Disease Transmission during an Influenza Pandemic

### 1 Lower and Upper Bound (LB and UB) Estimations for High-Risk Children and Adults

As all asthma patients are considered high-risk [1], we use a LB of 12% for children and LB of 8% for adults [2]. The UB is determined from our empirical data and arithmetic calculation. We find that 22% of the children are at high risks among those visiting the Children’s Healthcare of Atlanta (the largest provider of pediatric services in Georgia) in year 2009 and set it as the UB for children. We estimate 24% as the UB of adults who are high-risk (denoted as *P_hr_,_ad_*) based on (i) the high-risk proportion (denoted as *P_hr_,_overall_*, 20.7%=62/299 from a CDC report[3]) in the entire population, (ii) the proportions of adults and children in the population (denoted as *P_ad_,P_ch_* respectively), and (iii) the LB of high-risk children (denoted as *P_hr_,_ch_*). Specifically, the following is the formula we use to derive the UB of adults:

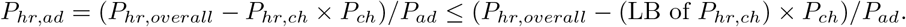

### 2 Natural Disease Progression Model and Parameters

We use a detailed SEIR (Susceptible, Exposed, Infectious, and Recovered) model [4, 5, 6] to depict the disease progression of individuals based on their ages and risk groups. In particular, individuals are divided into five age groups: 0–5, 6–11, 12–18, 19–64, 65+. They are either low-risk or high-risk. Each individual can stay in one of the disease stages, i.e. susceptible (*S*), exposed but not infectious (*E*), presymptomatic (*I_P_*), asymptomatic (*I_A_*), symptomatic (*I_S_*), hospitalized (*I_H_*), recovered (*R*) and dead (*D*), at a certain time. The disease progression model is shown in Figure 1 of Ekici, Keskinocak and Swann[5]. All individuals start from susceptible stage. If infected, individuals become exposed but not infectious and then presymptomatic. Presymptomatic patients may not show any symptom with probability *p_A_* or develop symptoms with probability 1 − *p_A_*. Asymptomatic patients recovers for sure. Symptomatic patients may be hospitalized with probability *p_H_* or directly get recovered with probability 1 − *p_H_*. Hospitalized patients may die with probability *p_D_* or recover with probability 1 − *p_D_*. Patients are infectious when they are in presymptomatic, asymptomatic, symptomatic and hospitalized stages. Individuals who get recovered are immune to the disease. The age-and-risk-specific parameters are presented in Table 1.

**Table 1.** Natural disease progression parameters.

| Notations | Descriptions | Values | References |
| --- | --- | --- | --- |
| $p_A$ | Probability of infected individual to become asymptomatic | 0.4 for working adults (age 19–24) and 0.25 for others | [4, 7, 8, 9] |
| $p_H$ | Probability of symptomatic individual to be hospitalized | Among low-risk people, 0.18 for young children (age 0–5), 0.12 for elderly (age 65+), and 0.06 for others; among high-risk people, 0.36 for young children (age 0–5), 0.12 for children (age 5–18) and elderly (age 65+), and 0.06 for others. | [4, 9] |
| $p_D$ | Probability of hospitalized individual who becomes dead | 0.344 for children (age 0–5) and elderly (age 65+), 0.172 for others | [4, 10] |
| Duration of $E + I_P$ | Length of exposed and presymptomatic stage | Weibull with mean 1.48 days and standard deviation 0.47 days and offset of 0.5 days | [4, 11] |
| Duration of $I_S$ | Length of symptomatic stage | Exponential with mean 2.7313 days | [4] |
| Duration of $I_A$ | Length of asymptomatic stage | Exponential with mean 1.63878 days | [4] |
| Duration of $I_H$ | Length of hospitalized stage | Exponential with mean 14 days | [4, 11] |
| Initial $R_0$ | Reproductive rate (average number of secondary cases generated by an infectious individual) before hospitals and clinics are introduced | 1.5, 1.8 | [4, 12, 9, 11, 13] |
| $\theta$ | Proportion of transmission that occurs at presymptomatic or asymptomatic stage | 0.3 | [4] |
| $\omega$ | Proportion of infections generated by individuals who are asymptomatic | 0.15 | [4] |
| $\gamma$ | Proportion of transmission that occurs outside the households | 0.7 | [12] |
| $\delta$ | Proportion of transmission outside the households that occurs in the community | 0.5 | [12] |

### 3 Contact Network and Parameters

In our simulation model, individuals contact others in social groups, including household (*H*), community (*C*), peer groups (*G*), hospitals (*D*), and flu clinics (*F*). Peer groups are classrooms for children and workplaces for adults. The average peer group sizes are 14, 20 and 30 for children in age groups 0–5, 6–11 and 12–18 respectively [14]. The workplace sizes for adults in age group 19–64 follow a truncated Poisson distribution with mean 20 and maximum 1000 [8]. Elderly with age 65 or above stay alone in his/her peer group. Initially, all individuals are susceptible. We randomly pick 30 individuals in the population to get infected.

Our simulation model determines the time of next infection and chooses a person to get infected based on the method of instantaneous FOI (Force Of Infection) in prior studies [4, 5, 6]. A higher instantaneous FOI experienced by a susceptible individual implies a higher probability the individual become infected in the social group. The instantaneous FOI experienced by the *i*th person in the day (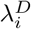) and in the night (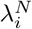) are calculated using the following formulas. Notations and calculations will be explained in details following the formulas.

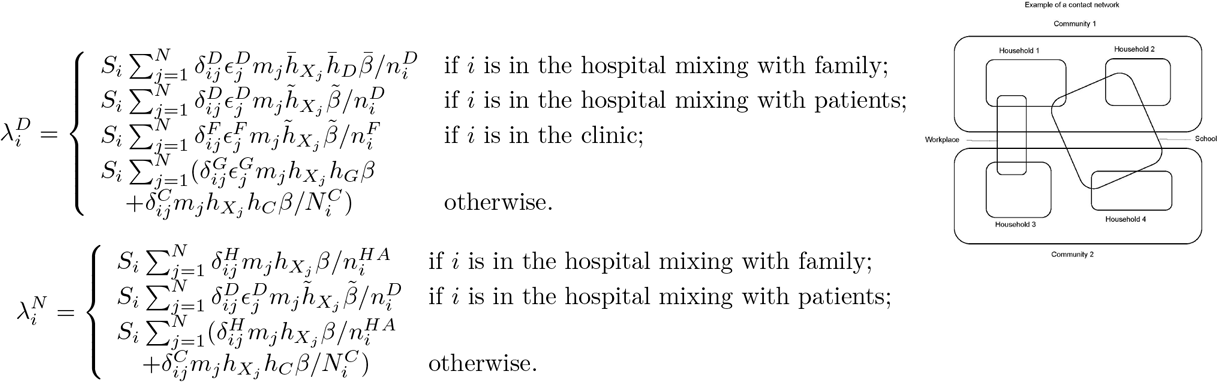

In particular, *S_i_* is the susceptibility of the *i*th individual and *m_i_* is the infectivity of a symptomatic individual *i*. They are defined the same as prior papers [5, 6, 10], i.e.,

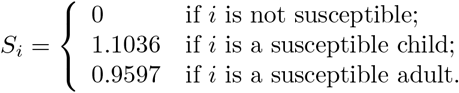

and

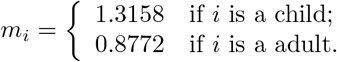

Moreover, 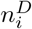 and 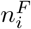 denote the number of individuals in the same hospital or clinic as the *i*th individual respectively. 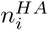 is the number of active household members of the ith individual, which excludes those dead and hospitalized individuals in *i*th individual’s household. 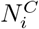 is the number of individuals in the *i*th individual’s community. 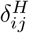, 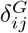, 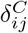, 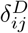 and 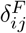 are indicators: if *i*th and *j*th individuals are within the same social group (household, peer group, community, hospital, and clinic respectively), the variable is set to be 1; otherwise the variable is 0. We assume that all symptomatic children withdraw from their peer groups and symptomatic adults withdraw from work with probability 0.5. 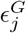, 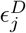 and 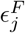 are indicators of *j*th individual being in his/her peer group, hospital and clinic respectively.

In our model, we consider three major maxing patterns: (i) The basic-mixing routine, where individuals mix in their peer groups during daytime, in their household groups during the night, and have some random contacts in their community groups, e.g. church and grocery stores, both day and night; (ii) The hospital/clinic-mixing routine and mixing with family mode at night, where individuals mix in hospitals and are accompanied by family members during the night; (iii) The hospital/clinic-mixing routine and mixing with patients mode at night, where individuals mix in hospitals/clinics and are not accompanied by family members during the night.

(i) is modeled in prior studies [4, 5, 6]. These studies define coefficient of transmission (*β*), relative hazards of an infected individual at disease stage *X* to symptomatic stage (*h_X_*, *X* in *{P,A,S*}), and relative hazards in social group *Y* to households (*h_Y_*, *Y* in *{H,G,C*}). Similarly to these definitions, we define 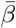, 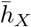 (*X* in {*P*, *A*, *S*}), 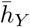 (*Y* in *{H,D*}) for (ii), and 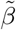, 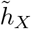 (*X* in *{P, A*, *S*}) for (iii). For (iii), 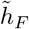 is defined to be one. Based on the definition, 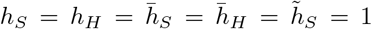. Note that we regard hospitalized individuals as symptomatic patients in hospitals so we will use *h_S_* (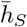 or 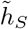) in the calculation of their infectivities. We will explain the details of estimating these parameters in three subsections.

Furthermore, 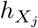, 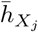 and 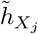 denote the relative hazard rate of the *j*th individual in disease stage *X* for (i), (ii) and (iii) respectively. We have the following relationships.

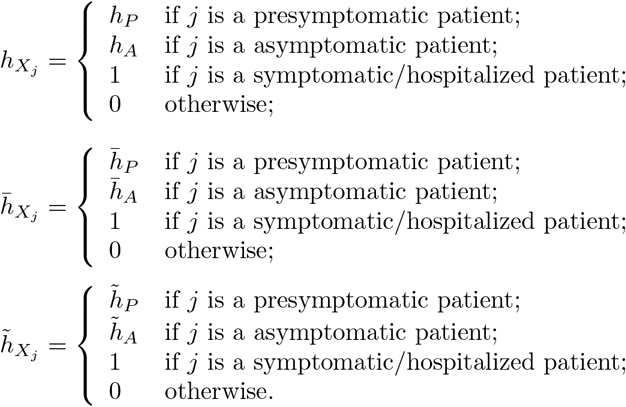

#### 3.1 Estimate of Parameters for Basic-Mixing Routine

In this subsection, we use initial *R*_0_ (reproduction number), *θ* (proportion of transmission that occurs at presymptomatic and asymptomatic stage), *ω* (proportion of infections generated by individuals who are asymptomatic), *γ* (proportion of transmission that occurs outside the households) and *δ* (proportion of transmission outside the households that occurs in the community) in Table 1 to estimate *β, h_X_* (*X* in *{P,A}*) and *h_Y_* (*Y* in *{G,C*}). The calibration method is used in prior studies [4, 5, 6]. We define *r_XY_* (X in *{P, A, S*} and *Y* in {*H, G, C*}) as the average number of people in social group *Y* by an individual at disease stage *X*. We can derive the following equations by definition.

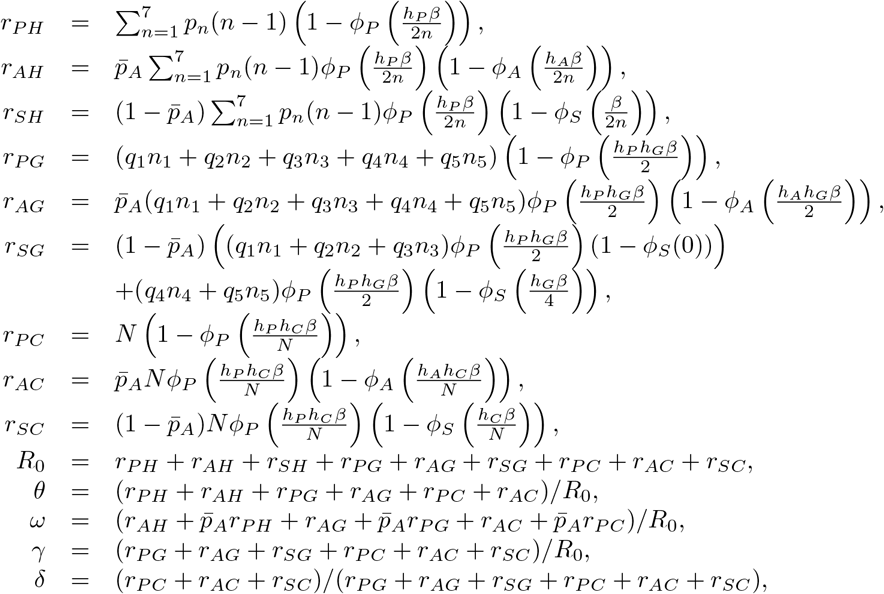

where *q_i_* denotes the proportion of population in age group *i* (*i* = 1,…, 5), *n_i_* is average size of peer groups for age group *i* (*i* = 1,…, 5), *N* is the total number of population, and 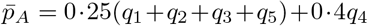, which is the average probability that a presymptomatic individual does not develop symptoms. *p_n_* (*n* = 1,…, 7) is the probability that an individual lives in a household with n members. [15] In addition, 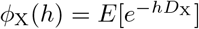 (*X* in *{P, A*, *S*}) defines the probability that an infection does not occur during disease stage *X* for a hazard of infection *h*, where the duration *D_X_* of disease stage *X* is defined in Table 1.

We solve the above nonlinear equations for *β*, *h_X_* (*X* in *{P, A*}) and *h_Y_* (*Y* in {*G*, *C*}).

#### 3.2 Estimate of Parameters for Hospital/Clinic-Mixing Routine and Mixing with Family at night

Similar to previous subsection, we can solve the following nonlinear equations for 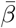, 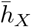 (*X* in *{P, A*}), 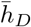.

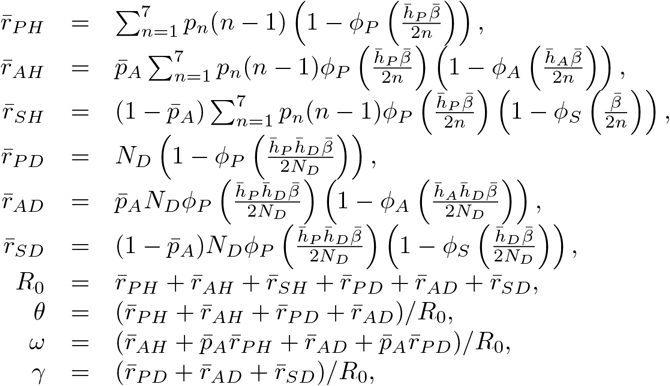

where 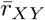 (*X* in {*P*, *A*, *S*} and *Y* in {*H*, *D*}) is defined as the average number of people in social group *Y* by an individual at disease stage *X* in mixing pattern (ii) and *N_D_* is the average number of patients in hospitals.

#### 3.3 Estimate of Parameters for Hospital/Clinic-Mixing Routine and Mixing with patients at night

Let 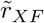 (*X* in *{P, A*, *S*}) be the average number of people in hospitals/clinics by an individual at disease stage *X* in mixing pattern (iii). Similar to subsections above, we can solve the following equations for 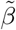, 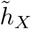 (*X* in *{P, A*}), and 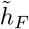.

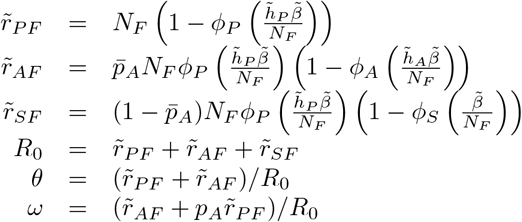

where *N_F_* is the average number of patients in hospitals/clinics.

### 4 Model Validation

We utilize the clinical attack rates for the 1957 pandemic [16] to validate our model. Clinical attack rate is the cumulative proportion of people who have ever been symptomatic. We adjust parameters as shown in Table 2. Table 3 presents the age-specific results of *R*_0_ =1.5, which is in line with the estimated *R*_0_ =1.5−1.7 in the 1957 pandemic [12]. The calibration procedure has been employed by several prior papers [5, 6, 9, 11, 17].

**Table 2.** Adjusted parameters to achieve the age-specific clinical attack rates for the 1957 pandemic.

| Parameter | Original | Adjusted |
| --- | --- | --- |
| $p_A$ | 0.4 for working adults (age 19–24) and 0.25 for others | 0.35 for age 0–18, 0.47 for age 19–64, 0.65 for age 65+ |
| $S_i$ | 1.1036 for susceptible children, 0.9597 for susceptible adults, 0 if not susceptible. | 1.4236 for susceptible children, 0.8374 for susceptible adults, 0 if not susceptible. |

**Table 3.**
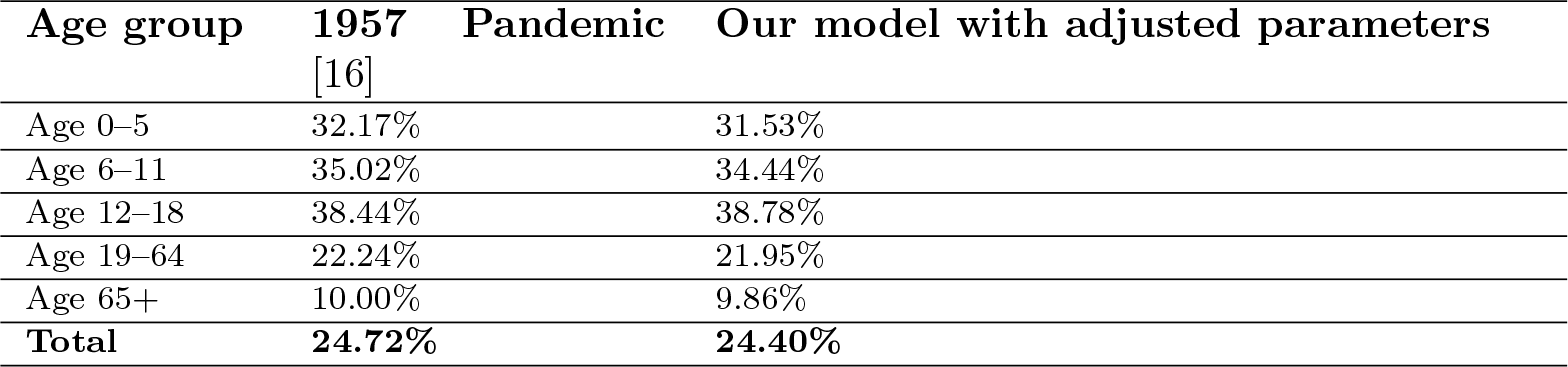
Age-specific clinical attack rates for our model validation.

## Notes

### Competing Interest Statement

The authors have declared no competing interest.

